# Altered transcriptomes, cell type proportions, and dendritic spine morphology in hippocampus of suicide deaths

**DOI:** 10.1101/2023.01.28.23285121

**Authors:** Sujan C. Das, Anton Schulmann, William B. Callor, Leslie Jerominski, Mitradas M. Panicker, Erik D. Christensen, William E. Bunney, Megan E. Williams, Hilary Coon, Marquis P. Vawter

## Abstract

Suicide is a condition resulting from complex environmental and genetic risks that affect millions of people globally. Both structural and functional studies identified the hippocampus as one of the vulnerable brain regions contributing to suicide risk. Here, we have identified the hippocampal transcriptomes, gene ontology, cell type proportions, dendritic spine morphology, and transcriptomic signature in iPSC-derived neuronal precursor cells (NPCs) and neurons in postmortem brain tissue from suicide deaths. The hippocampal tissue transcriptomic data revealed that *NPAS4* gene expression was downregulated while *ALDH1A2, NAAA*, and *MLXIPL* gene expressions were upregulated in tissue from suicide deaths. The gene ontology identified 29 significant pathways including *NPAS4*-associated gene ontology terms “excitatory post-synaptic potential”, “regulation of postsynaptic membrane potential” and “long-term memory” indicating alteration of glutamatergic synapses in the hippocampus of suicide deaths. The cell type deconvolution identified decreased excitatory neuron proportion and an increased inhibitory neuron proportion providing evidence of excitation/inhibition imbalance in the hippocampus of suicide deaths. In addition, suicide deaths had increased dendric spine density, due to an increase of thin (relatively unstable) dendritic spines, compared to controls. The transcriptomes of iPSC-derived hippocampal-like NPCs and neurons revealed 31 and 33 differentially expressed genes in NPC and neurons, respectively, of suicide deaths. The suicide-associated differentially expressed genes in NPCs were *RELN, CRH, EMX2, OXTR, PARM1* and *IFITM2* which overlapped with previously published results. The previously-known suicide-associated differentially expressed genes in differentiated neurons were *COL1A1, THBS1, IFITM2, AQP1*, and *NLRP2*. Together, these findings would help better understand the hippocampal neurobiology of suicide for identifying therapeutic targets to prevent suicide.

## Introduction

Suicide is a complex and polygenic disorder affecting millions of people worldwide [1]. In 2020, suicide took the lives of ∼46,000 people in the U.S. making it one of the top 9 leading causes of death for people aged 10-64 in the USA (Centers for Disease Control and Prevention, 2020). Suicide is a consequence of complex interactions between stressors (external/internal) and suicide-related traits [2]. Memory deficits are one of the suicide-related traits frequently associated with suicide risk [2]. Suicide attempters show poor cognition and memory performances, which are among the strongest cognitive risk factors for suicide deaths [3-5]. Poor social functioning history was also observed in suicide deaths [6]. Structural neuroimaging studies showed that both suicide attempters and deaths had decreased hippocampal volume, possibly related to the cognitive deficits associated with suicide risks [7-9]. In addition to abnormal hippocampal structures, suicide attempters also had increased brain activity and connectivity in bilateral hippocampus [10-12]. Thus, strong evidence implicates the hippocampus as a significant brain region involved in the pathophysiology of suicide.

Human hippocampus is involved in memory and cognition. The human hippocampus consists of pharmacologically and functionally distinct sub-regions such as the cornu ammonis (CA1-4), the dentate gyrus (DG), and the subiculum. Different sub-regions of the hippocampus have been shown to respond differently in previous studies of suicidality and suicide deaths. For example, suicide attempters with strong familial risk showed decreased volumes of CA1 and CA3 [13]. Suicide deaths showed decreased DG volume and number of granule neurons in DG [7]. A hippocampal transcriptomics study of suicide deaths showed altered expression of genes involved in glutamatergic neurotransmission, synaptic plasticity, and cell adhesion, among others [14,15]. Genome-wide association studies (GWAS) of suicide attempts also revealed glutamatergic synapses as a significant pathway in suicide attempters across multiple ancestries [16]. Since different sub-regions of the hippocampus responds differently in suicide outcomes, a region-specific transcriptomic study is required to identify the subregion-specific hippocampal neuropathology associated with suicide risk. In addition, synapse-level structural alteration in sub-regions of the hippocampus tissue from suicide deaths will potentially identify how alterations in glutamatergic synapses is involved in risk of suicide death. Moreover, given that suicide is about 35-48% heritable [17,18], the induced pluripotent stem cell (iPSC)-derived neural precursor cells (NPCs) and neurons from suicide deaths could show similar suicide-associated transcriptional signatures, reflecting underlying genetic liability rather than acquired differences due to external non-genetic exposures.

The present study aims to fill critical gaps in our current understanding of the hippocampal neuropathology of suicide, with converging evidence from several synergistic experiments. First, we have identified differentially expressed genes and significant gene ontology terms in the CA1 sub-region of the hippocampus of suicide deaths with next-generation sequencing (RNA-Seq). In order to identify suicide-associated changes in relative cell type proportion, we have determined the estimated cell type proportion using Bisque [19] within the CA1 region of suicide deaths. To identify how changes in the glutamatergic synapse structures are associated with suicide risk, we have analyzed the morphology of high-resolution and three-dimensional dendritic spines of CA1 pyramidal neurons in tissues from suicide deaths. Finally, we have identified the differentially expressed genes in iPSC-derived hippocampal-like neural precursor cells (NPCs) and neurons from suicide deaths. To our best knowledge, this is the first study showing dendritic spine-level alterations and altered transcriptomes in iPSC-derived hippocampal-like NPCs and neurons of suicide deaths. These robust results in iPSC-derived neurons open a window of opportunity for future large-scale studies in research cohorts where postmortem brain samples may not be available.

## Materials and Methods

### Samples

Postmortem hippocampal CA1 tissues from 50 deceased subjects and skin biopsies from 4 of those 50 subjects were used for this study. Out of these 50 subjects, 42 postmortem hippocampal tissues and 4 skin biopsies were obtained through the Utah State Office of the Medical Examiner (OME). The brain and skin biopsy sample collection at the University of Utah were approved under an exemption (45CFR46.102(f)) from the Institutional Review Board (IRB). Four (4) brain tissues were obtained from the NIH Neurobiobank. The remaining 4 postmortem hippocampal tissues were obtained through the University of California Irvine (UCI). The postmortem brain collection at the UCI was also approved by the UCI Institutional Review Board (IRB). The study had two experimental groups: non-suicide controls (CON) (n=28) and suicide deaths (Sui) (n=22). The full description of the demographics of these subjects is presented in Table ST1A and ST1B.

### RNA Extraction and Sequencing

The details of the RNA extraction and sequencing are described in the supplementary information.

### Gene-level Differential Expression (DE) and Gene Ontology (GO) Analyses

The differentially expressed genes (DEGs) analysis was performed using DESeq2. The gene ontology (GO) analysis was performed with the gene score resampling (GSR) method of the ErmineJ [20]. The details of these procedures are described in the supplementary information.

### qPCR

The relative mRNA expression of *NPAS4* was determined through quantitative PCR (qPCR). Briefly, cDNA was produced with TaqMan Reverse Transcription Reagent (Invitrogen N8080234). The qPCR was carried out in triplicate with QuantStudio 7 Flex System (Applied Biosystems) in reaction mixture containing PowerUp SYBR Green Master Mix (Applied Biosystems, A25776), respective primers (*NPAS4* or *GAPDH*) and cDNA. The relative mRNA expression of *NPAS4* was quantified using the 2^-∆∆CT^ method.

### Cell Types Deconvolution

Deconvolution was performed via Bisque [19] using the reference data-based method for seven major cell types: excitatory neurons, inhibitory neurons, astrocytes, oligodendrocytes, oligodendrocyte precursor cells (OPC), microglia/T-cells, and mural cells. The raw gene count matrix from the bulk RNA-Seq data was used as input data. Published single-nucleus RNA-Seq data from postmortem human hippocampus [21] served as the reference data. Single-nucleus RNA-seq data was visualized via a UMAP plot based on the top 20 principal components in Seurat.

### Dendritic Spine Morphology Analyses

Dendritic spine morphology of CA1 pyramidal neurons was analyzed as described as our previously published method [22]. Please see the supplementary information for the details.

### Conversion of Skin Fibroblasts into iPSC

Generation of iPSCs from dermal fibroblasts (from 2 control subjects and 2 suicide deaths) was performed based on methods described previously [23,24]. For the detailed procedure, please see the supplementary information.

### Generation of Hippocampal-like Neural Precursor Cell (NPC) and Neurons

Hippocampal-like NPCs and neurons were obtained from iPSC as described previously with slight modification [25,26]. The detailed procedure has been described in the supplementary information.

### Statistical Analyses

The differentially expressed genes (DEGs) between two groups (CON vs Sui) was carried out using DESeq2 with default parameters which uses the Wald test, and the obtained p-values are corrected for multiple testing using the Benjamini and Hochberg method [27]. The cell type deconvolution coefficients (estimated cell type proportion) were analyzed by linear regression with group (CON, Sui), age, sex, and RIN as covariates for individual cell type. The two group comparisons for dendritic spine density, spine length and spine diameter were conducted by mixed model analyses using group (CON, Sui) with age, sex, and PMI as independent variables. The dendritic spine type (thin, stubby, mushroom) by dendrite type (apical, basal) analysis was performed with a mixed model incorporating spine type, dendrite type, age, sex, PMI, and group as independent variables using the repeated measure function in JMP v16.1 (SAS Institute).

## Results

### Differentially Expressed Genes (DEGs) and Gene Ontology (GO) in CA1 of Suicide Deaths

CA1 tissues from 28 controls and 22 suicide deaths were included in differential gene expression analysis. The detailed information about the demographics of the subjects has been depicted in Table ST1A and Table ST1B. The age and RNA Integrity Number (RIN) was not significantly different between CON and Sui groups (Table ST1A). However, postmortem interval (PMI) was significantly higher (p=0.04) in Sui group compared to CON (Table ST1A). DESeq2 (∼age+sex+RIN+group) revealed four (4) significant differentially expressed genes (DEGs) in CA1 of suicide deaths (Sui) compared to controls (CON) with FDR≤0.5 and p≤0.05 (Figure 1). The 4 DEGs were Neuronal PAS Domain Protein 4 (*NPAS4*), Aldehyde Dehydrogenase 1 Family Member A2 (*ALDH1A2*), N-Acylethanolamine Acid Amidase (*NAAA*) and (MLX Interacting Protein Like) (*MLXIPL*). *NPAS4* [p=8.98E-06, FDR=0.03, FC=5.81, LSMean (CON)=175.70, LSMean (Sui)=30.26] was significantly down-regulated while *ALDH1A2* [p=5.59E-07, FDR=0.01, FC=4.16, LSMean (CON)=51.56, LSMean (Sui)=214.69], *NAAA* [p=4.72E-06, FDR=0.03, FC=1.32, LSMean (CON)=89.16, LSMean (Sui)=117.35] and *MLXIPL* [p=8.39E-06, FDR=0.03, FC=1.59, LSMean (CON)=55.89, LSMean (Sui)=88.68] was significantly upregulated in suicide deaths (Sui) compared to controls (CON). qPCR further validated the decreased relative gene expression of NPAS4 in Sui group compared to CON group (Figure 1).

**Figure 1:**
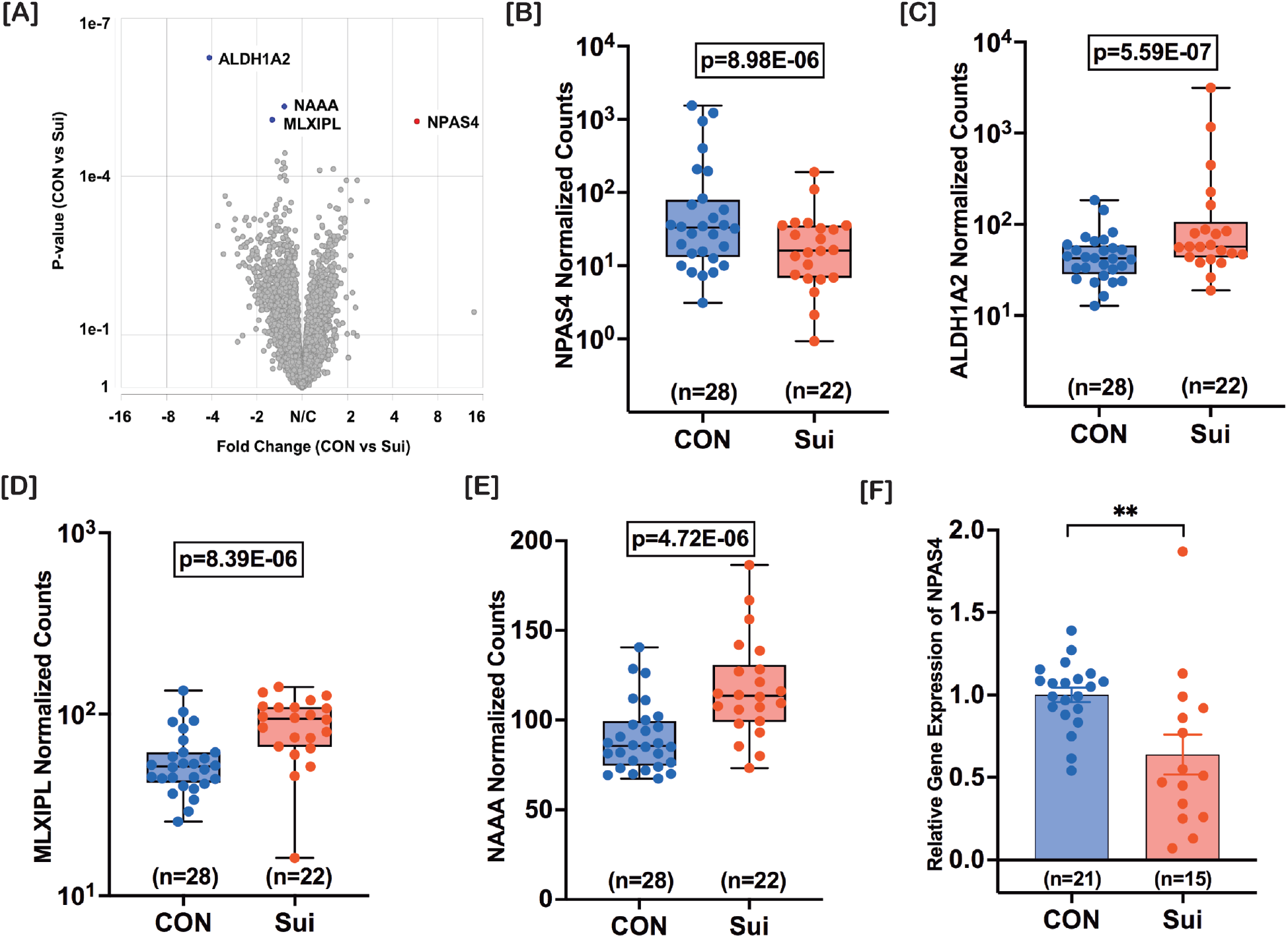
Differentially expressed genes (DEGs) in hippocampal CA1 region of suicide deaths. The DEGs were analyzed through DESeq2. The volcano-plot shows *NPAS4, ALDH1A2, NAAA* and *MLXIPL* as DEGs in suicide deaths (CON vs Sui, FDR≤0.05, p≤0.05) [A]. The *NPAS4* gene expression was significantly downregulated in suicide deaths (FC=5.81, p=8.98E-06) [B] while *ALDH1A2* (FC=4.16, p=5.59E-07) [C], *MLXIPL* (FC=1.59, p=8.39E-06) [D] and *NAAA* (FC=1.32, p=4.72E-06) [E] gene expression was significantly upregulated in suicide group. qPCR validated the decreased relative gene expression of *NPAS4* in the suicide deaths [F]. CON=control group; Sui=suicide group; FC=fold change; FDR=false discovery rate; **, p<0.01.

GO analysis using the gene score resampling (GSR) method implemented in ErmineJ (version 3.2) revealed 29 significant biological processes (GO terms) at corrected p<0.05 and corrected multifunctionality p<0.05 (Table ST2). *NPAS4*-associated significant GO terms include “excitatory postsynaptic potential”, “long-term memory”, and “regulation of postsynaptic membrane potential” which are suggestive of alteration in excitatory postsynaptic potential in CA1 neurons of suicide deaths. The details of gene ontology term “GO:0060079-Excitatory Postsynaptic Potential” is depicted in Figure SF3. There were 13 significant GO terms associated with *ALDH1A2* including “vitamin A metabolic process”, “response to retinoic acid”, “cellular response to retinoic acid”, and “retinoic acid biosynthetic process”. The “primary alcohol metabolic process” GO term was associated for both *ALDH1A2* and *NAAA*. There were no significant GO terms associated for *MLXIPL*. Of interest, the top 3 significant GO terms were related to interferon production which are suggestive of alerted immune-responses in suicide deaths (Table ST2).

### Estimate Cell Type Proportions in Hippocampal CA1 of Suicide deaths

The estimated cell type proportions with hippocampal CA1 were determined for the seven major cell types: excitatory neurons, inhibitory neurons, astrocytes, oligodendrocytes, oligodendrocytes progenitor cells (OPC), microglia/T-cells, and mural cells (Figure SF1). The linear regression analysis identified significant group (CON vs Sui) effects for the estimated cell proportion of excitatory and inhibitory neurons. The estimated cell type proportion of excitatory neurons was significantly higher (p=0.03) while inhibitory neuron proportion was significantly lower (p=0.004) in suicide deaths compared to controls (Figure 2). The group effects for astrocytes (p=0.13), oligodendrocytes (p=0.69), OPC (p=0.63), mural cells (p=0.96), and microglia/T-cell (p=0.24) were not statically significant (Figure 2). The inhibitory neuron proportion negatively correlated with age (p=0.02) while females had significantly (p=0.008) lower excitatory neuron proportions compared to their male counterparts (Figure SF2).

**Figure 2:**
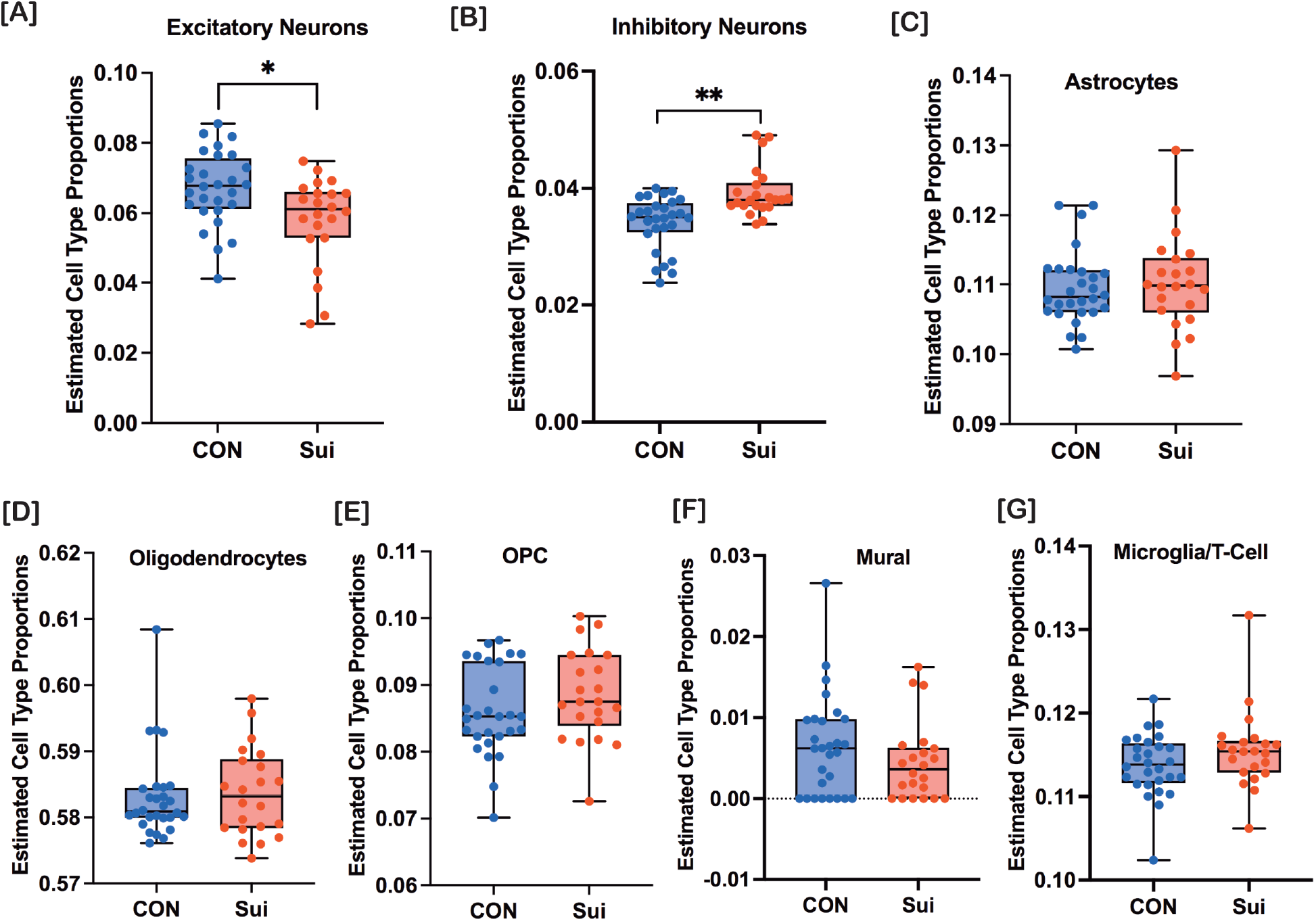
Estimated cell type proportions in hippocampal CA1 of suicide deaths. The cell type deconvolution was performed using Bisque with the raw bulk RNA-Seq data with published single-nucleus RNA-Seq data from human hippocampus as a reference. Linear regression analysis revealed that the estimated excitatory neuron proportion [A] was significantly higher while inhibitory neuron proportion [B] was significantly lower in suicide deaths (Sui) compared to controls (CON). The estimated cell type proportion for astrocytes [C], oligodendrocytes [D], OPC [E], mural cells [F], and microglia/T-cell [G] was not significantly different between controls and suicide deaths. OPC=Oligodendrocyte Progenitor Cells, CON=Controls, Sui=Suicide deaths. n=28 for CON, n=22 for Sui. *, p<0.05; **, p<0.01.

### Dendritic Spine Morphology of CA1 Pyramidal Neurons of Suicide Deaths

Neuroplasticity-related genes have been shown to downregulate in the hippocampus of suicide deaths [28]. Thus, we hypothesized that suicide affects the glutamatergic synapses at their structural level. We conducted dendritic spine morphology analyses in CA1 pyramidal neurons to test this hypothesis. Both the apical dendritic spine density (LSMean of CON=2.33, Std. Error=0.141; LSMean of Sui=3.15, Std. Error=0.264; p=0.015) and the basal dendritic spine density (LSMean of CON=2.00, Std. Error=0.134; LSMean of Sui=2.897, Std. Error=0.251; p=0.00511) were significantly higher in suicide deaths (Sui) compared to controls (CON) (Figure 3). The increased spine density in suicide deaths was associated with significantly decreased apical spine head width (LSMean of CON=0.397, Std. Error=0.0053; LSMean of Sui=0.358, Std. Error=0.011; p=0.004) and unaltered basal spine head width (LSMean of CON=0.394, Std. Error=0.0058; LSMean of Sui=0.364, Std. Error=0.016; p=0.10) in suicide deaths. However, the spine lengths of apical (p=0.71) and basal (p=0.15) spines were not significantly different between controls and suicide deaths. We further analyzed the spine types (thin, stubby and mushroom) to determine which spine type contributed to the increased spine density. A two-way repeated measure analysis of spine type x dendrite type revealed significant group x spine type interaction (p=0.00008). The increased spine density (both apical and basal) was due to increase in thin spines in both apical dendrites (p=1.3E-6) and basal (p=2.9E-7) dendrites (Figure 3). The stubby and mushroom type dendritic spines were not significantly different between controls and suicide deaths (Figure 3). Age, sex, and PMI did not have any significant effect on any of the dendritic spine parameters. These results are in line with our previous findings where we showed that age (up to 71 years-old) and PMI (up to 28 hours) did not have statistically significant effects on CA1 dendritic spine morphology parameters (i.e., spine density, spine head diameter, spine length) of adult subjects [22].

**Figure 3:**
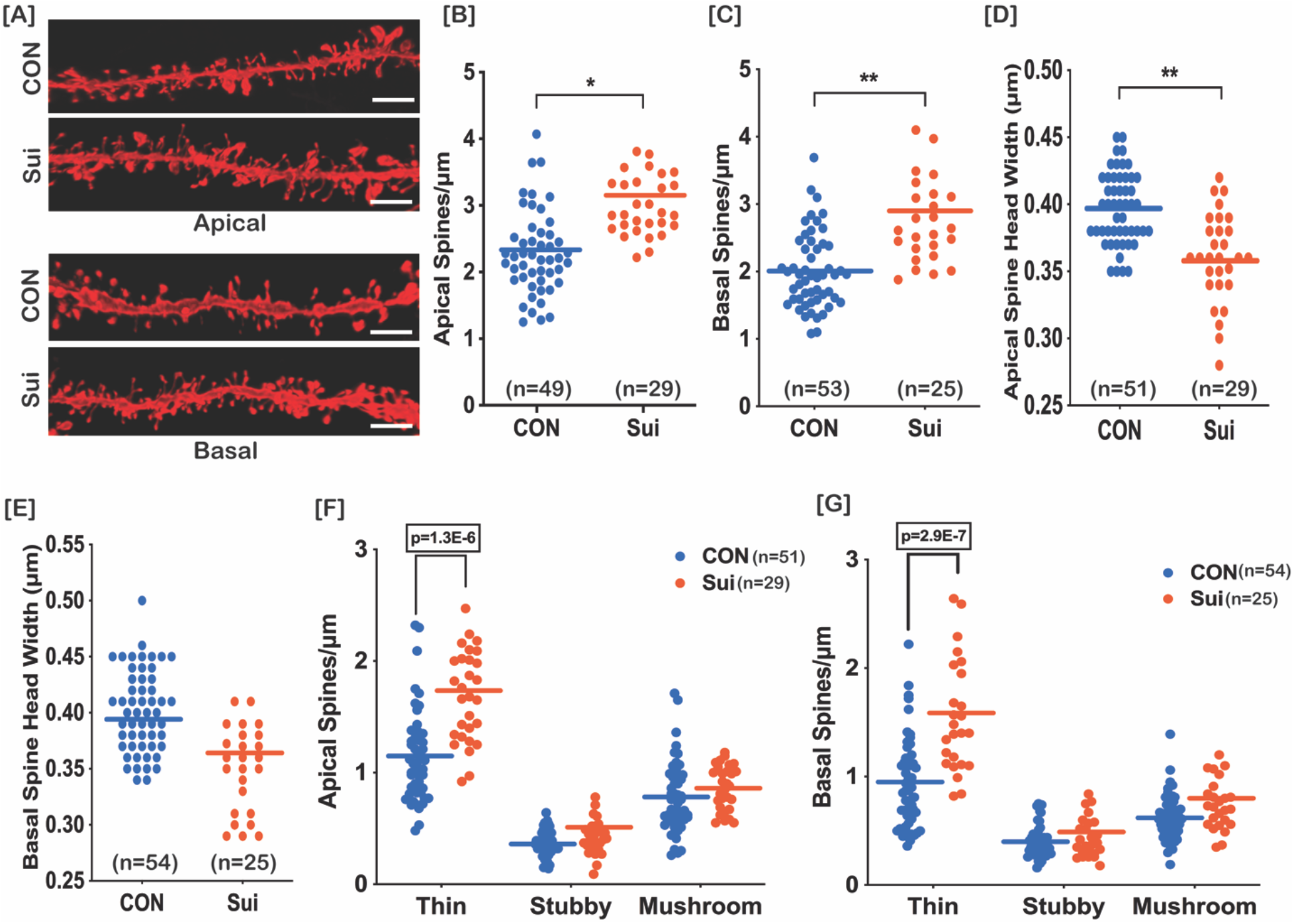
Dendritic spine morphology of hippocampal CA1 pyramidal neurons of suicide deaths. The 3D deconvolved apical (upper panel) and basal (lower panel) dendritic segments from control and suicide deaths were modeled and analyzed using Neurolucida 360 [A]. Both apical [B] and basal [C] dendritic spine density were significantly higher in suicide deaths compared to controls. The increased apical and basal spine densities were associated with decreased apical spine head width [D] and unaltered basal spine head width [E] in suicide deaths. The increased apical and basal dendritic spine density was due to an increase of thin dendritic spines in both apical [F] and basal [G] dendrites. The stubby and mushroom spines were not significantly different in suicide deaths compared to controls. Horizontal lines=Least Square Mean (LSM), CON=control group; Sui=suicide group; n=number of neurons (2-3 dendritic segments/neuron). Scale bar = 4 µm. *, p<0.05; **, p<0.01.

### Differentially Expressed Genes (DEGs) in iPSC-derived hippocampal-like NPCs and Neurons of Suicide Deaths

Postmortem tissues serve as a tremendous resource for identifying genetic and molecular markers for suicide. Unfortunately, postmortem tissues constrain the functional studies crucial for identifying suicide-associated functional abnormalities of neurons. Because suicide is about 35-48% heritable, we hypothesized that iPSC-derived NPCs and neurons from suicide deaths would show similar suicide-associated transcriptomic signatures, reflecting underlying genetic liability rather than acquired differences due to external non-genetic exposures. As a pilot study, we have generated iPSCs from fibroblasts of 2 suicide deaths and 2 controls. Hippocampal-like NPCs and neurons were derived from the corresponding iPSCs. The iPSC-derived hippocampal-like neurons were characterized through bulk RNA-Seq and immunostaining. The transcriptomic profile in both NPCs and neurons of suicide deaths were determined through bulk RNA-Seq. Five weeks in-vitro (5WIV) differentiated neurons showed, by RNA-Seq, enriched expression of neuronal marker genes such as *MAP2, TUBB3* and *DLG4* (Table ST3). The differentiated neurons had enriched expression of *ZBTB20, LHX9, ELAVL2* and *TSPAN7* which are known hippocampus marker genes [26,29,30] (Table ST3). Differential gene expression analysis via DESeq2 revealed upregulation of 15 known markers for rodent and human CA3 (and CA2) in differentiated neurons (vs NPCs) (Table ST3). Known CA1 markers (POU3F1, WFS1) were not significantly enriched in 5 WIV differentiated neurons (vs NPC) (Table ST3). The expression of neuronal markers MAP2 and beta-tubulin-III were also confirmed through immunocytochemistry (ICC) (Figure 4). The majority of cells (5 WIV) were glutamatergic as ∼78% (% of DAPI) of cells were positive for glutamatergic neuron marker vGluT1. The differentiated neurons also expressed excitatory neuron marker PSD95 at 6 weeks in-vitro (6 WIV). The number of GABA-positive cells was ∼7-10% (Figure 4). DG, CA1 and cortical neuron maker CTIP2 [31] was not consistently expressed. Approximately 67% cells (% of DAPI) were positive for CA3 marker GRIK4 whereas ∼35% cells were positive for DG marker PROX1 (Figure 4).

**Figure 4:**
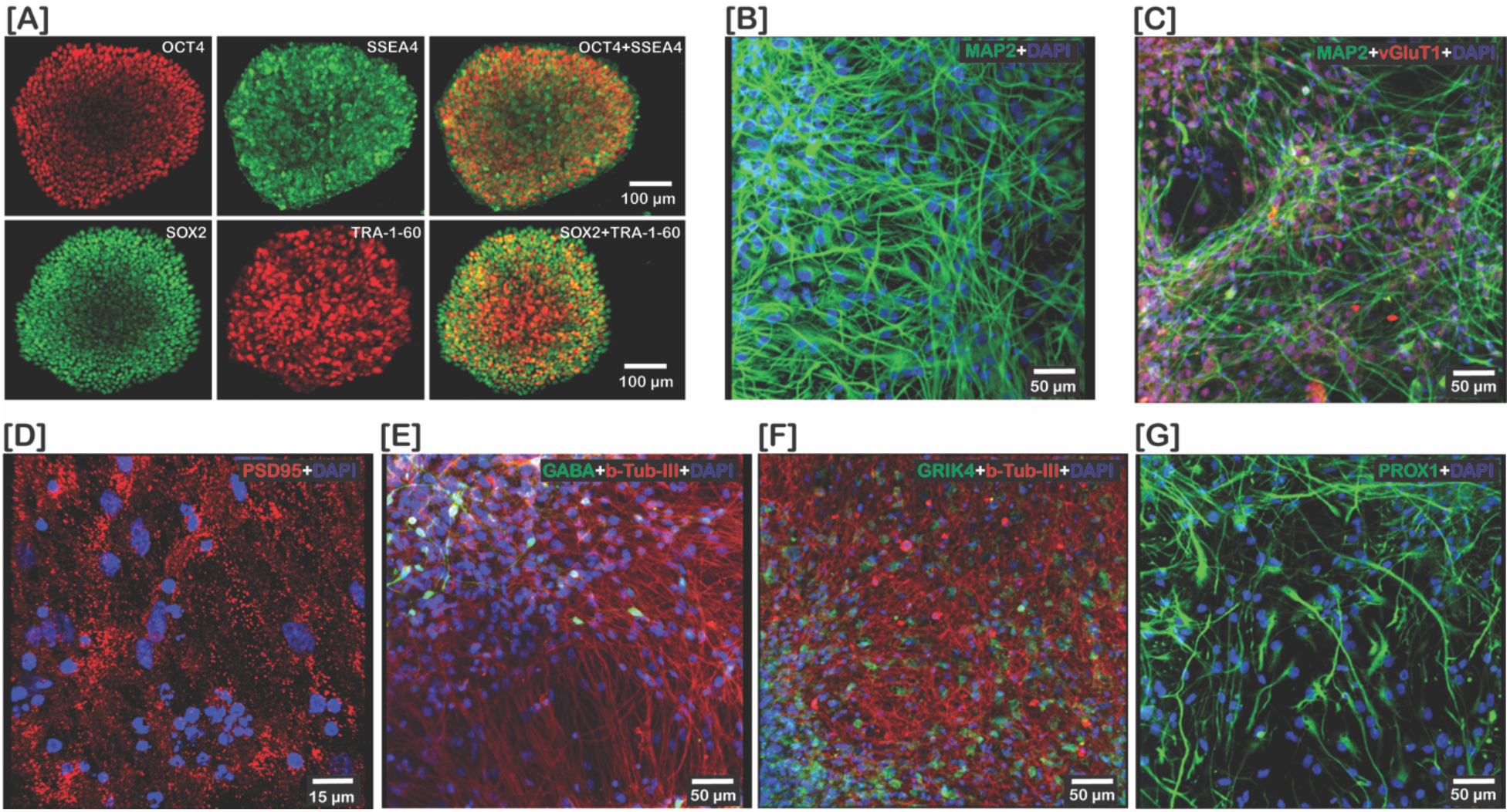
Characterization of iPSCs and iPSC-derived neurons. The skin fibroblast-derived iPSCs were positive for pluripotency markers OCT4, SOX2, SSEA4 and TRA-1-60 [A]. The iPSC-derived neurons were positive for neuronal marker MAP2 [B] and beta-Tubulin-III [E]. The majority (∼78%) of cells were positive for glutamatergic neuron marker vGluT1 [C]. The differentiated neurons also expressed synaptic marker PSD95 [D]. About 7-10% cells were positive for GABA [E]. The CA3 marker GRIK4 was positive for ∼67% cells [F] whereas ∼35% cells also expressed DG marker PROX1 [G].

Next, we determined the DEGs in NPC and differentiated neurons of suicide deaths through DESeq2 (vs CON). DESeq2 (CON vs Sui; study design: ∼age+group) identified 31 and 33 differentially expressed genes (DEGs) (p≤0.05, FDR≤0.05) in iPSC-derived NPCs and neurons, respectively, of suicide deaths (Table 1). The details of 31 DEGs in iPSC-derived NPCs of suicide deaths is shown in Table ST4. The details of 33 DEGs in iPSC-derived neurons of suicide deaths is shown in Table ST5. For iPSC-derived NPCs, the already-known suicide-associated DEGs are *RELN* (Reelin), *CRH* (Corticotropin Releasing Hormone), *EMX2* (Empty Spiracles Homeobox 2), *OXTR* (Oxytocin Receptor), *PARM1* (Prostate Androgen-Regulated Mucin-Like Protein 1), and *IFITM2* (Interferon Induced Transmembrane Protein 2). For iPSC-derived neurons, previously-known suicide associated DEGs are *COL1A1* (Collagen Type I Alpha 1 Chain), *THBS1* (Thrombospondin 1), *IFITM2* (Interferon Induced Transmembrane Protein 2), *AQP1* (Aquaporin 1), and *NLRP2* (NLR Family Pyrin Domain Containing 2).

**Table 1:** Differentially expressed genes (DEGs), through RNA-Sequencing, in iPSC-derived hippocampal-like NPCs and neurons of suicide deaths. n=2/group. There are 31 and 33 DEGs in hippocampal-like NPCs and neurons, respectively, of suicide deaths. p≤0.05, FDR≤0.05. Green-highlighted and yellow-highlighted genes are overlapping genes with prior evidence from studies of suicide and depressive disorder, respectively.

| DESeq2 results for iPSC-derived hippocampal-like NPCs (CON vs Sui) |  |  |  |  |  |
| --- | --- | --- | --- | --- | --- |
| Up-regulated in Suicide |  |  |  | Down-regulated in Suicide |  |
| *SNORD3B-2* | *CCZ1B* |  |  | *RELN* | *OXTR* |
| *SNORD3B-1* | *COL12A1* |  |  | *CRH* | *COL8A1* |
| *H4C11* |  |  |  | *CNTN2* | *CCZ1* |
| *COL3A1* |  |  |  | *SCUBE1* | *BLACAT1* |
| *HMCN1* |  |  |  | *EMX2OS* | *PRSS23* |
| *PDPR* |  |  |  | *PARM1* | *ABCA1* |
| *H2AC12* |  |  |  | *TMTC1* | *FZD5* |
| *BGN* |  |  |  | *CNTNAP2* | *UNC5D* |
| *FBLN2* |  |  |  | *EMX2* | *VAV3* |
| *IFITM2* |  |  |  | *SCG2* |  |
| DESeq2 results for iPSC-derived hippocampal-like Neurons (CON vs Sui) |  |  |  |  |  |
| Up-regulated in Suicide |  |  |  | Down-regulated in Suicide |  |
| *SNORD3B-2* | *COL6A3* | *FBN1* | *NLRP2* | *TBR1* |  |
| *SNORD3B-1* | *H4C11* | *MME* | *MXRA5* |  |  |
| *COL3A1* | *PRRX1* | *MMP2* |  |  |  |
| *COL1A1* | *SNORD97* | *PDGFRA* |  |  |  |
| *DCN* | *IFITM2* | *MGP* |  |  |  |
| *IGFBP3* | *FBN2* | *ALDH1A3* |  |  |  |
| *COL5A1* | *ID1* | *COL5A2* |  |  |  |
| *HMCN1* | *POSTN* | *AQP1* |  |  |  |
| *THBS1* | *CYP26A1* | *CCZ1B* |  |  |  |
| *LUM* | *NOMO3* | *CXCL14* |  |  |  |

## Discussion

### Transcriptomes, Cell Type Proportion, and Gene Ontology in Hippocampal CA1 are Altered in Suicide

The hippocampal CA1 transcriptomics identified four differentially expressed genes: *NPAS4, ALDH1A2, MLXIPL*, and *NAAA*. Among these four genes, *NPAS4* had the highest fold change in suicide deaths (fold change=5.81, p=8.98E-06) (Figure 1). Neuronal PAS Domain Protein 4 (*NPAS4*) is an immediate-early gene transcription factor and exclusively expressed in neurons. In hippocampal neurons, *NAPS4* maintains the synaptic excitation and inhibition balance by the development of inhibitory synapses on excitatory neurons through regulating activity-dependent gene expression [32]. *NPAS4* also interacts with neuronal activity-dependent promoters to mediate the expression of brain-derived neurotrophic factor (BDNF) [33]. *NPAS4* plays a crucial role in contextual memory formation through the regulation of hippocampal CA3 transcriptional program [34]. *NPAS4* also regulates neurite outgrowth and offers neuroprotection from excitotoxic neurodegeneration [35]. Thus, decreased *NPAS4* expression in suicide deaths might have the following risk-related implications: (a) alteration of the balance between excitation and inhibition; (b) increased memory deficits; (c) alteration of the structural and functional synaptic plasticity.

Biological pathway affected by the NPAS4 were also significantly associated with suicide deaths. There were three *NPAS4*-associated significant gene ontology terms among which the most significant gene ontology term was “excitatory postsynaptic potential” (Table ST2). This result confirms that changes in the glutamatergic synapses within the hippocampus may be a significant risk factor for suicide death. A recent large GWAS study which also identified glutamatergic synapses as an altered pathway in suicide attempters [16]. Thus, glutamatergic synapses might serve as druggable targets to prevent suicide. Since decreased NPAS4 is an indication of imbalance in neuronal excitation and inhibition, we further explored possible alterations in the relative proportions of major cell types. Interestingly, the relative proportion of excitatory neurons was lower while the relative proportion of inhibitory neurons was higher in suicide deaths. Suicide attempters have been shown to have smaller hippocampus [8,36]. Thus, decreased excitatory neuron proportions might be linked to the decreased hippocampal volumes observed in suicidality. A very recent study found that the relative proportion of nine major cell types were not significantly different in DLPFC of suicide deaths compared to non-psychiatric controls [37]. This discrepancy might be attributed to brain-regional differences or other differences in ascertainment.

### Increased Thin Dendritic Spines in CA1 of Suicide Deaths

Dendritic spines are small protrusions arising from the neuronal dendritic shaft and serve as morphological specialization (post-synaptic compartments) for excitatory synapses. The majority of excitatory synapses end in dendritic spines [38]. Thus, the number of dendritic spines (spine density) positively correlate with the number of excitatory synapses. Moreover, the shape of the dendritic spines relates the maturity (synaptic strength) of the excitatory synapses (maturity in decreasing order: mushroom>stubby>thin) [38]. The mushroom-shaped dendritic spines represent the matured synapses while thin-shaped dendritic spines represent immature synapses (called “learning synapses”). We found that there was an increase in dendritic spine density, driven by increase in thin dendritic spines, in CA1 neurons of suicide deaths. Dendritic spines are plastic in nature and spine categories can switch types. Therefore, there is a possibility that the increased thin dendritic spines in suicide deaths might be due to the conversion of stubby and mushroom spines into thin spines. However, there was no change in the number of stubby and mushroom dendritic spines. Thus, the increased thin dendritic spines are more likely to be newly formed immature spines in suicide deaths. Similar synaptic instability (increase in unstable spines) has been reported in a rodent model of depressive disorders. Chronic social stress, a rodent model of depression, increased immature stubby dendritic spines in accumbal medium spiny neurons [39]. The FDA-approved antidepressant ketamine has been shown to alleviate depressive symptoms through increasing mushroom (matured) dendritic spines [40]. Thus, targeting synaptogenesis or increasing mushroom dendritic spines might be an effective strategy to alleviate depressive disorder and/or suicide.

### iPSC-derived hippocampal-like NPCs and Neurons from Suicide Deaths Show Previously-known Suicide-associated Gene Expression Signatures

In this pilot study, there were 15 genes (26%) out of 57 DEGs (in both NPC and neurons) that overlapped with prior differential gene expression studies in suicide and/or depressive disorder (Table 1). The six genes with overlapping evidence from previous studies of suicide and depression that were downregulated in NPCs of our suicide death cohort were *CRH, RELN, EMX2, PARM1, OXTR*, and *CNTNAP2*. The corticotropin releasing hormone (*CRH*) gene and protein expression has been shown to be significantly increased in prefrontal cortex of teenage suicide deaths [41,42] and epigenetic changes in *CRH* gene has been associated with increased risk for suicidality [43]. The relative expression of Reelin (*RELN*) was significantly decreased in the locus coeruleus of suicide deaths [44]. In addition, hippocampal dentate gyrus *RELN* expression was decreased in depressed subjects [45]. *EMX2* gene expression was significantly down-regulated in the prefrontal cortex of suicide deaths with schizophrenia [46]. The *PARM1* gene expression was upregulated in the hippocampus of suicide deaths [15]. Oxytocin is a neuropeptide which plays a crucial role in complex social behavior. The effects of oxytocin depends on the concentration of its receptor called OXTR. Oxytocin levels and OXTR expression have been associated with suicidality and depressive disorders. For example, suicide attempters showed decreased levels of oxytocin in blood and cerebrospinal fluid (CSF) [47-49]. A single nucleotide polymorphism (SNP) within the *OXTR* gene has been associated with suicide attempt [50], and OXTR expression was significantly higher in prefrontal cortex of subjects with major depressive disorder (MDD) [51]. CNTNAP2 mediates cell-cell interactions in the brain and a SNP in this gene has been shown to be associated with major depression [52].

Three overlapping genes that were upregulated in NPCs of our suicide deaths were *IFITM2, HMCN1*, and *FBLN2*. Results for *IFITM2* and *HMCN1* were found for both NPCs and neurons (Table 1). The IFITM2 gene has been shown to be downregulated in dorsolateral prefrontal cortex of female suicide deaths compared to female controls [53]. FBLN2 activates the TGF-beta1 signaling pathway. Micro RNA miR-192-5p, which targets the *FBLN2* gene, was found to be upregulated in synaptic fractions of dorsolateral prefrontal cortex of subjects with major depression [54]. In addition, suppressing FBLN2 expression has been shown to rescue the cognitive impairment in a mouse model of depression [55]. The six overlapping genes that were upregulated in our iPSC-derived neurons of suicide deaths were *COL1A1, THBS1, AQP1, NLRP2, IGFBP3*, and *MMP2*. Among them, we previously showed that functional sequence variants within *COL1A1* has been shown to pose a significant familial risk of suicide deaths [56]. The synaptic-related gene *THBS1* was differentially expressed in female depressed mice [57,58], and a recent proteomic study found that THBS1 protein expression was upregulated in DLPFC of suicide deaths [59]. Reduced expression of the *AQP1* gene was reported in the dorsomedial prefrontal cortex of suicide deaths [60]. NLRP is one of a family of NOD-like receptors containing pyrin, which are innate immunity receptors that play a crucial role to form the inflammasome in brain. We found that *NLRP2* was significantly increased in iPSC-derived hippocampal-like neurons of suicide deaths. NLRP1, NLRP3 and NLRP6 protein and mRNA expression was found to be increased significantly in prefrontal cortex of suicide deaths [61]. Cerebrospinal fluid (CSF) level of MMP2 was significantly higher in patients with major depression [62] while peripheral levels of MMP2 were significantly lower in patients with mood disorders [63]. A recent study showed that iPSC-derived NPCs and neurons from MDD patients had decreased basal respiration and increased spontaneous electrical activity, respectively [64]. iPSC-derived GABA interneurons from suicide attempters with major depression showed increased neurite branches, electrophysiological hyperexcitability, and decreased calcium signaling [65]. Thus, iPSC-derived NPC and neuron cultures from suicide deaths might serve as an important, scalable model for suicide-associated neuropathology.

### Conclusions

In this study, we have identified decreased gene expression of *NPAS4* and increased gene expression of *ALDH1A2, MLXIPL* and *NAAA* in the hippocampus of suicide deaths. These altered transcriptomes are associated with decreased excitatory cell proportion and increased inhibitory cell proportion. In addition, there is an increase in thin dendritic spines in hippocampal pyramidal neurons of suicide deaths. The iPSC-derived hippocampal-like NPCs and neurons from suicide deaths implicated previously-known suicide-associated transcriptomic signatures. Together, these findings will help to better understand the hippocampal neuropathology of suicide. Importantly, the use of iPSC-derived neuron cultures from suicide deaths might serve as a scalable “disease in-vitro” model of suicide neuropathology.

## Data Availability

The RNA-Seq raw data will be deposited to NCBI Gene Expression Omnibus (GEO).

## Data Availability

The RNA-Seq raw data will be deposited to NCBI Gene Expression Omnibus (GEO).

## Acknowledgments

We would like to thank staff at the Utah State Office of the Medical Examiner (OME) for their assistance in acquiring skin biopsies and brain tissues. We are grateful to the NIH Neurobiobank for providing brain tissues.

## Authors Contributions

Drafted and edited manuscript (all). Designed and supervised study (HC, MPV). Wrote manuscript (SCD, HC, MPV). Sample collection (WBC, EDC, SCD, WEB, LJ). Conducted laboratory experiments (SCD, MMP, MEW). Data analysis (SCD, AS, MPV).

## Funding and Disclosure

National Institute of Mental Health [R01MH122412 and R01MH123489 to HC] [R01MH085801 to MPV]; American Foundation for Suicide Prevention (AFSP) [to SCD, 2019 Pilot Research Grant (AFSP Grant ID# PRG-1-032-18)]; University of California Della Martin Foundation [to SCD, Postdoctoral Fellowship]. Postmortem brain collection at the University of California-Irvine (UCI) Brain Bank was supported by the Pritzker Neuropsychiatric Disorders Research Consortium.

## Competing Interests

The authors have nothing to disclose Data Sharing

The RNA-Seq raw data will be deposited to NCBI Gene Expression Omnibus (GEO).

## Notes

### Competing Interest Statement

The authors have declared no competing interest.

### Author Declarations

The brain and skin biopsy sample collection at the University of Utah were approved under an exemption (45CFR46.102(f)) from the Institutional Review Board (IRB). The postmortem brain collection at the UCI was also approved by the UCI Institutional Review Board (IRB).

## References

1 Sisti D, Mann JJ, Oquendo MA. Toward a Distinct Mental Disorder-Suicidal Behavior. JAMA Psychiatry. 2020;77(7):661–62.

2 Mann JJ, Rizk MM. A Brain-Centric Model of Suicidal Behavior. Am J Psychiatry. 2020;177(10):902–16.

3 Zhang L, Lucassen PJ, Salta E, Verhaert P, Swaab DF. Hippocampal neuropathology in suicide: Gaps in our knowledge and opportunities for a breakthrough. Neurosci Biobehav Rev. 2022;132:542–52.

4 Huber RS, Sheth C, Renshaw PF, Yurgelun-Todd DA, McGlade EC. Suicide Ideation and Neurocognition Among 9- and 10-Year Old Children in the Adolescent Brain Cognitive Development (ABCD) Study. Arch Suicide Res. 2020:1–15.

5 Sinyor M, Williams M, Mitchell R, Zaheer R, Bryan CJ, Schaffer A, et al. Cognitive behavioral therapy for suicide prevention in youth admitted to hospital following an episode of self-harm: A pilot randomized controlled trial. J Affect Disord. 2020;266:686–94.

6 Weiser M, Fenchel D, Werbeloff N, Goldberg S, Fruchter E, Reichenberg A, et al. The association between premorbid cognitive ability and social functioning and suicide among young men: A historical-prospective cohort study. Eur Neuropsychopharmacol. 2017;27(1):1–7.

7 Boldrini M, Galfalvy H, Dwork AJ, Rosoklija GB, Trencevska-Ivanovska I, Pavlovski G, et al. Resilience Is Associated With Larger Dentate Gyrus, While Suicide Decedents With Major Depressive Disorder Have Fewer Granule Neurons. Biol Psychiatry. 2019;85(10):850–62.

8 Colle R, Chupin M, Cury C, Vandendrie C, Gressier F, Hardy P, et al. Depressed suicide attempters have smaller hippocampus than depressed patients without suicide attempts. J Psychiatr Res. 2015;61:13–8.

9 Chen F, Bertelsen AB, Holm IE, Nyengaard JR, Rosenberg R, Dorph-Petersen KA. Hippocampal volume and cell number in depression, schizophrenia, and suicide subjects. Brain Res. 2020;1727:146546.

10 Wagner G, Li M, Sacchet MD, Richard-Devantoy S, Turecki G, Bar KJ, et al. Functional network alterations differently associated with suicidal ideas and acts in depressed patients: an indirect support to the transition model. Transl Psychiatry. 2021;11(1):100.

11 Weng JC, Chou YS, Tsai YH, Lee CT, Hsieh MH, Chen VC. Connectome Analysis of Brain Functional Network Alterations in Depressive Patients with Suicidal Attempt. J Clin Med. 2019;8(11).

12 Lan MJ, Rizk MM, Pantazatos SP, Rubin-Falcone H, Miller JM, Sublette ME, et al. Restingstate amplitude of low-frequency fluctuation is associated with suicidal ideation. Depress Anxiety. 2019;36(5):433–41.

13 Pantazatos S, Ogden, T., Melhem, N., Allsop, D. B., Lesanpezeshki, M., Burke, A., … Mann, J. J. (2022, February 17). Smaller volumes of the cornu ammonis (CA1 and CA3) may be a biological endophenotype for suicide risk in mood disorders. https://doi.org/10.31234/osf.io/x9p6s. x2022.

14 Kouter K, Zupanc T, Videtic Paska A. Genome-wide DNA methylation in suicide victims revealing impact on gene expression. J Affect Disord. 2019;253:419–25.

15 Glavan D, Gheorman V, Gresita A, Hermann DM, Udristoiu I, Popa-Wagner A. Identification of transcriptome alterations in the prefrontal cortex, hippocampus, amygdala and hippocampus of suicide victims. Sci Rep. 2021;11(1):18853.

16 Kimbrel NA, Ashley-Koch AE, Qin XJ, Lindquist JH, Garrett ME, Dennis MF, et al. A genomewide association study of suicide attempts in the million veterans program identifies evidence of pan-ancestry and ancestry-specific risk loci. Mol Psychiatry. 2022;27(4):2264–72.

17 Otsuka I, Akiyama M, Shirakawa O, Okazaki S, Momozawa Y, Kamatani Y, et al. Genome-wide association studies identify polygenic effects for completed suicide in the Japanese population. Neuropsychopharmacology. 2019;44(12):2119–24.

18 Docherty AR, Shabalin AA, DiBlasi E, Monson E, Mullins N, Adkins DE, et al. Genome-Wide Association Study of Suicide Death and Polygenic Prediction of Clinical Antecedents. Am J Psychiatry. 2020;177(10):917–27.

19 Jew B, Alvarez M, Rahmani E, Miao Z, Ko A, Garske KM, et al. Accurate estimation of cell composition in bulk expression through robust integration of single-cell information. Nat Commun. 2020;11(1):1971.

20 Gillis J, Mistry M, Pavlidis P. Gene function analysis in complex data sets using ErmineJ. Nat Protoc. 2010;5(6):1148–59.

21 Tran MN, Maynard KR, Spangler A, Huuki LA, Montgomery KD, Sadashivaiah V, et al. Single-nucleus transcriptome analysis reveals cell-type-specific molecular signatures across reward circuitry in the human brain. Neuron. 2021;109(19):3088–103 e5.

22 Das SC, Chen D, Callor WB, Christensen E, Coon H, Williams ME. DiI-mediated analysis of presynaptic and postsynaptic structures in human postmortem brain tissue. J Comp Neurol. 2019;527(18):3087–98.

23 Fujimori K, Tezuka T, Ishiura H, Mitsui J, Doi K, Yoshimura J, et al. Modeling neurological diseases with induced pluripotent cells reprogrammed from immortalized lymphoblastoid cell lines. Mol Brain. 2016;9(1):88.

24 Okita K, Matsumura Y, Sato Y, Okada A, Morizane A, Okamoto S, et al. A more efficient method to generate integration-free human iPS cells. Nat Methods. 2011;8(5):409–12.

25 Pomeshchik Y, Klementieva O, Gil J, Martinsson I, Hansen MG, de Vries T, et al. Human iPSC-Derived Hippocampal Spheroids: An Innovative Tool for Stratifying Alzheimer Disease Patient-Specific Cellular Phenotypes and Developing Therapies. Stem Cell Reports. 2020;15(1):256–73.

26 Sarkar A, Mei A, Paquola ACM, Stern S, Bardy C, Klug JR, et al. Efficient Generation of CA3 Neurons from Human Pluripotent Stem Cells Enables Modeling of Hippocampal Connectivity In Vitro. Cell Stem Cell. 2018;22(5):684–97 e9.

27 Love MI, Huber W, Anders S. Moderated estimation of fold change and dispersion for RNA-seq data with DESeq2. Genome Biol. 2014;15(12):550.

28 Fuchsova B, Alvarez Julia A, Rizavi HS, Frasch AC, Pandey GN. Altered expression of neuroplasticity-related genes in the brain of depressed suicides. Neuroscience. 2015;299:1–17.

29 Abellan A, Desfilis E, Medina L. Combinatorial expression of Lef1, Lhx2, Lhx5, Lhx9, Lmo3, Lmo4, and Prox1 helps to identify comparable subdivisions in the developing hippocampal formation of mouse and chicken. Front Neuroanat. 2014;8:59.

30 Sakaguchi H, Kadoshima T, Soen M, Narii N, Ishida Y, Ohgushi M, et al. Generation of functional hippocampal neurons from self-organizing human embryonic stem cell-derived dorsomedial telencephalic tissue. Nat Commun. 2015;6:8896.

31 Williams ME, Wilke SA, Daggett A, Davis E, Otto S, Ravi D, et al. Cadherin-9 regulates synapse-specific differentiation in the developing hippocampus. Neuron. 2011;71(4):640–55.

32 Lin Y, Bloodgood BL, Hauser JL, Lapan AD, Koon AC, Kim TK, et al. Activity-dependent regulation of inhibitory synapse development by Npas4. Nature. 2008;455(7217):1198–204.

33 Yoshihara S, Takahashi H, Nishimura N, Kinoshita M, Asahina R, Kitsuki M, et al. Npas4 regulates Mdm2 and thus Dcx in experience-dependent dendritic spine development of newborn olfactory bulb interneurons. Cell Rep. 2014;8(3):843–57.

34 Ramamoorthi K, Fropf R, Belfort GM, Fitzmaurice HL, McKinney RM, Neve RL, et al. Npas4 regulates a transcriptional program in CA3 required for contextual memory formation. Science. 2011;334(6063):1669–75.

35 Woitecki AM, Muller JA, van Loo KM, Sowade RF, Becker AJ, Schoch S. Identification of Synaptotagmin 10 as Effector of NPAS4-Mediated Protection from Excitotoxic Neurodegeneration. J Neurosci. 2016;36(9):2561–70.

36 Gosnell SN, Velasquez KM, Molfese DL, Molfese PJ, Madan A, Fowler JC, et al. Prefrontal cortex, temporal cortex, and hippocampus volume are affected in suicidal psychiatric patients. Psychiatry Res Neuroimaging. 2016;256:50–56.

37 Punzi G, Ursini G, Chen Q, Radulescu E, Tao R, Huuki LA, et al. Genetics and Brain Transcriptomics of Completed Suicide. Am J Psychiatry. 2022;179(3):226–41.

38 Hering H, Sheng M. Dendritic spines: structure, dynamics and regulation. Nat Rev Neurosci. 2001;2(12):880–8.

39 Golden SA, Christoffel DJ, Heshmati M, Hodes GE, Magida J, Davis K, et al. Epigenetic regulation of RAC1 induces synaptic remodeling in stress disorders and depression. Nat Med. 2013;19(3):337–44.

40 Li N, Lee B, Liu RJ, Banasr M, Dwyer JM, Iwata M, et al. mTOR-dependent synapse formation underlies the rapid antidepressant effects of NMDA antagonists. Science. 2010;329(5994):959–64.

41 Pandey GN, Rizavi HS, Bhaumik R, Ren X. Increased protein and mRNA expression of corticotropin-releasing factor (CRF), decreased CRF receptors and CRF binding protein in specific postmortem brain areas of teenage suicide subjects. Psychoneuroendocrinology. 2019;106:233–43.

42 Merali Z, Kent P, Du L, Hrdina P, Palkovits M, Faludi G, et al. Corticotropin-releasing hormone, arginine vasopressin, gastrin-releasing peptide, and neuromedin B alterations in stress-relevant brain regions of suicides and control subjects. Biol Psychiatry. 2006;59(7):594–602.

43 Jokinen J, Bostrom AE, Dadfar A, Ciuculete DM, Chatzittofis A, Asberg M, et al. Epigenetic Changes in the CRH Gene are Related to Severity of Suicide Attempt and a General Psychiatric Risk Score in Adolescents. EBioMedicine. 2018;27:123–33.

44 Roy B, Wang Q, Palkovits M, Faludi G, Dwivedi Y. Altered miRNA expression network in locus coeruleus of depressed suicide subjects. Sci Rep. 2017;7(1):4387.

45 Knable MB, Barci BM, Webster MJ, Meador-Woodruff J, Torrey EF, Stanley Neuropathology C. Molecular abnormalities of the hippocampus in severe psychiatric illness: postmortem findings from the Stanley Neuropathology Consortium. Mol Psychiatry. 2004;9(6):609-20, 544.

46 Kim S, Choi KH, Baykiz AF, Gershenfeld HK. Suicide candidate genes associated with bipolar disorder and schizophrenia: an exploratory gene expression profiling analysis of post-mortem prefrontal cortex. BMC Genomics. 2007;8:413.

47 Chu C, Hammock EAD, Joiner TE. Unextracted plasma oxytocin levels decrease following in-laboratory social exclusion in young adults with a suicide attempt history. J Psychiatr Res. 2020;121:173–81.

48 Jokinen J, Chatzittofis A, Hellstrom C, Nordstrom P, Uvnas-Moberg K, Asberg M. Low CSF oxytocin reflects high intent in suicide attempters. Psychoneuroendocrinology. 2012;37(4):482–90.

49 Jahangard L, Shayganfard M, Ghiasi F, Salehi I, Haghighi M, Ahmadpanah M, et al. Serum oxytocin concentrations in current and recent suicide survivors are lower than in healthy controls. J Psychiatr Res. 2020;128:75–82.

50 Parris MS, Grunebaum MF, Galfalvy HC, Andronikashvili A, Burke AK, Yin H, et al. Attempted suicide and oxytocin-related gene polymorphisms. J Affect Disord. 2018;238:62–68.

51 Lee MR, Sheskier MB, Farokhnia M, Feng N, Marenco S, Lipska BK, et al. Oxytocin receptor mRNA expression in dorsolateral prefrontal cortex in major psychiatric disorders: A human post-mortem study. Psychoneuroendocrinology. 2018;96:143–47.

52 Ji W, Li T, Pan Y, Tao H, Ju K, Wen Z, et al. CNTNAP2 is significantly associated with schizophrenia and major depression in the Han Chinese population. Psychiatry Res. 2013;207(3):225–8.

53 Cabrera-Mendoza B, Fresno C, Monroy-Jaramillo N, Fries GR, Walss-Bass C, Glahn DC, et al. Sex differences in brain gene expression among suicide completers. J Affect Disord. 2020;267:67–77.

54 Yoshino Y, Roy B, Dwivedi Y. Differential and unique patterns of synaptic miRNA expression in dorsolateral prefrontal cortex of depressed subjects. Neuropsychopharmacology. 2021;46(5):900–10.

55 Tang CZ, Yang JT, Liu QH, Wang YR, Wang WS. Up-regulated miR-192-5p expression rescues cognitive impairment and restores neural function in mice with depression via the Fbln2-mediated TGF-beta1 signaling pathway. FASEB J. 2019;33(1):606–18.

56 Coon H, Darlington T, Pimentel R, Smith KR, Huff CD, Hu H, et al. Genetic risk factors in two Utah pedigrees at high risk for suicide. Transl Psychiatry. 2013;3:e325.

57 Seney ML, Logan RW. Critical roles for developmental hormones and genetic sex in stress-induced transcriptional changes associated with depression. Neuropsychopharmacology. 2021;46(1):221–22.

58 Paden W, Barko K, Puralewski R, Cahill KM, Huo Z, Shelton MA, et al. Sex differences in adult mood and in stress-induced transcriptional coherence across mesocorticolimbic circuitry. Transl Psychiatry. 2020;10(1):59.

59 Kim MJ, Do M, Han D, Son M, Shin D, Yeo I, et al. Proteomic profiling of postmortem prefrontal cortex tissue of suicide completers. Transl Psychiatry. 2022;12(1):142.

60 Dora F, Renner E, Keller D, Palkovits M, Dobolyi A. Transcriptome Profiling of the Dorsomedial Prefrontal Cortex in Suicide Victims. Int J Mol Sci. 2022;23(13).

61 Pandey GN, Zhang H, Sharma A, Ren X. Innate immunity receptors in depression and suicide: upregulated NOD-like receptors containing pyrin (NLRPs) and hyperactive inflammasomes in the postmortem brains of people who were depressed and died by suicide. J Psychiatry Neurosci. 2021;46(5):E538–E47.

62 Lindqvist D, Janelidze S, Erhardt S, Traskman-Bendz L, Engstrom G, Brundin L. CSF biomarkers in suicide attempters--a principal component analysis. Acta Psychiatr Scand. 2011;124(1):52–61.

63 Shibasaki C, Takebayashi M, Itagaki K, Abe H, Kajitani N, Okada-Tsuchioka M, et al. Altered Serum Levels of Matrix Metalloproteinase-2, -9 in Response to Electroconvulsive Therapy for Mood Disorders. Int J Neuropsychopharmacol. 2016;19(9).

64 Triebelhorn J, Cardon I, Kuffner K, Bader S, Jahner T, Meindl K, et al. Induced neural progenitor cells and iPS-neurons from major depressive disorder patients show altered bioenergetics and electrophysiological properties. Mol Psychiatry. 2022.

65 Lu K, Hong Y, Tao M, Shen L, Zheng Z, Fang K, et al. Depressive patient-derived GABA interneurons reveal abnormal neural activity associated with HTR2C. EMBO Mol Med. 2022:e16364.

