## Supplementary Information for "Altered transcriptomes, cell type proportions, and dendritic spine morphology in hippocampus of suicide deaths"

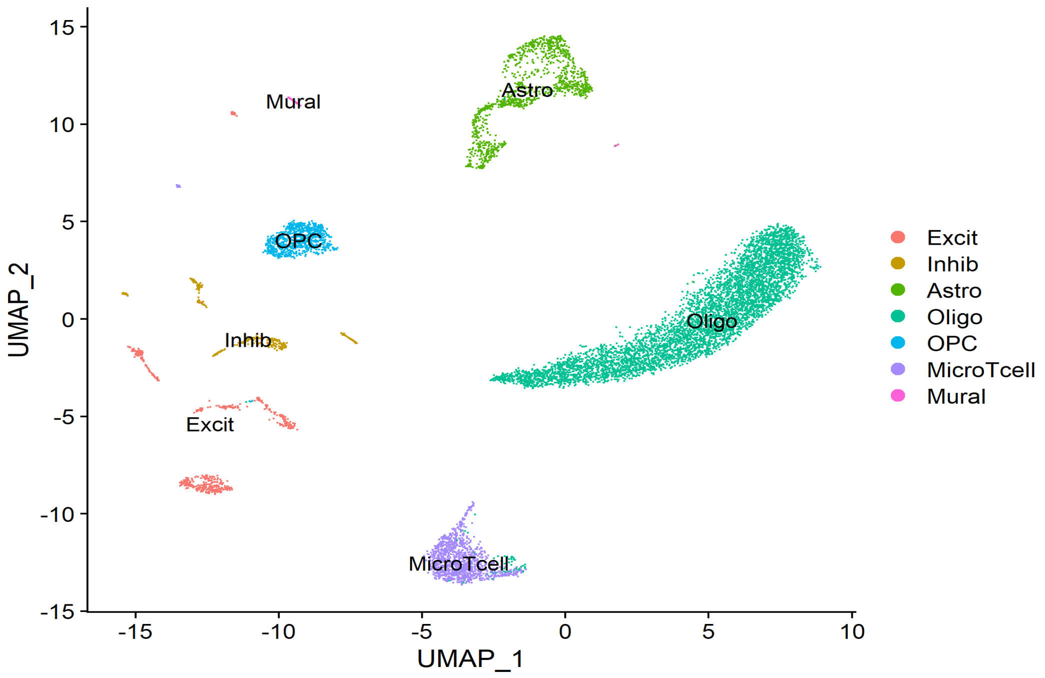

Figure SF1: Major cell types for which the estimated cell type proportions were determined. Excit=Excitatory Neurons, Inhib=Inhibitory Neurons, Astro=Astrocytes, Oligo=Oligodendrocytes, OPC=Oligodendrocytes Progenitor Cells.

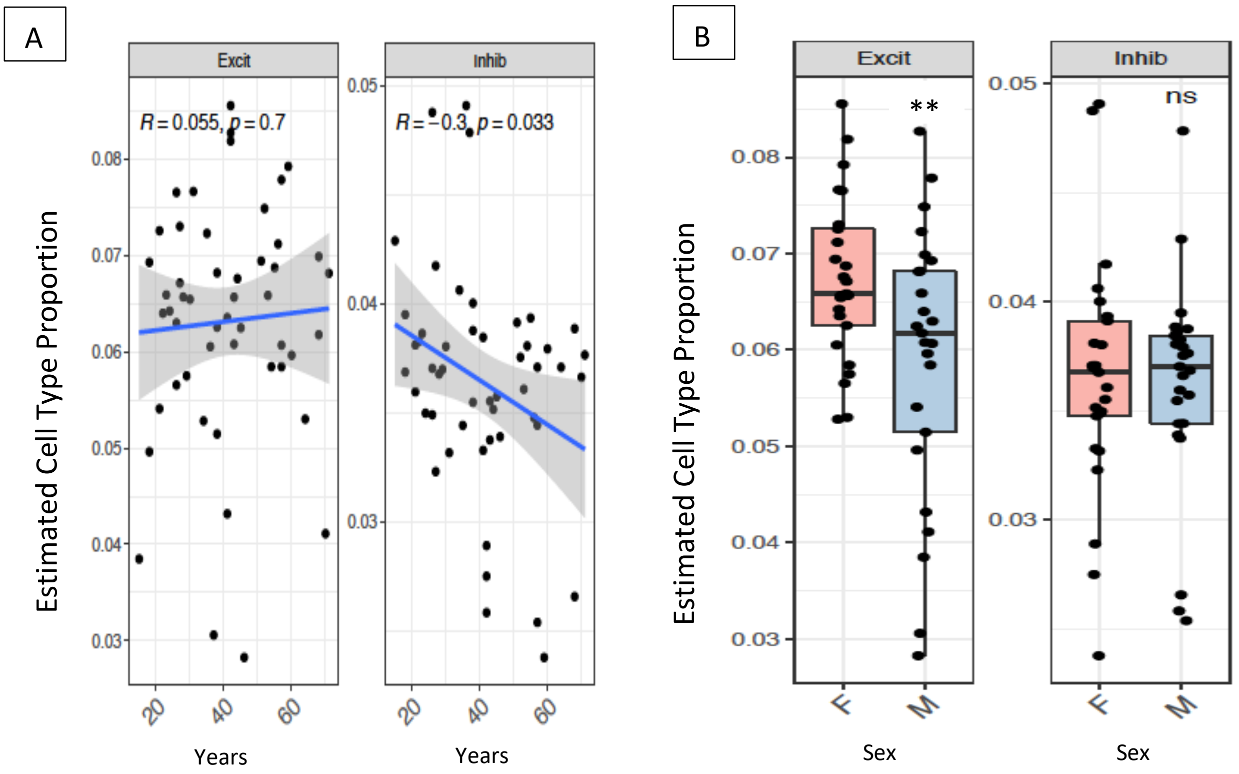

Figure SF2: The effect of age (A) and sex (B) on estimated cell type proportions of excitatory and inhibitory neurons. Excit=excitatory neurons, Inhib= inhibitory neurons, F=females, M=males.

GO:0060079 - excitatory postsynaptic potential (49 genes) Scores: DESeq2_Final.txt col: 2 pvalue

Cell Width: Color Range:

| Element | Score | QQ Score | Symbol | Name | Multifunc | QQ Multifunc |
|---|---|---|---|---|---|---|
| NPAS4 | 8.98e-06 | | NPAS4 | neuronal PAS domain protein 4 | 0.745 (131) | |
| P2RX6 | 5.48e-05 | | P2RX6 | purinergic receptor P2X 6 | 0.687 (109) | |
| CHRNA1 | 0.000642 | | CHRNA1 | cholinergic receptor nicotinic alpha 1 subunit | 0.805 (116) | |
| MEF2C | 0.00159 | | MEF2C | myocyte enhancer factor 2C | 0.997 (484) | |
| GLRA2 | 0.00566 | | GLRA2 | glycine receptor alpha 2 | 0.893 (126) | |
| GRIN2C | 0.00954 | | GRIN2C | glutamate ionotropic receptor NMDA type subunit 2C | 0.826 (176) | |
| P2RX4 | 0.0233 | | P2RX4 | purinergic receptor P2X 4 | 0.953 (298) | |
| CHRNB1 | 0.0513 | | CHRNB1 | cholinergic receptor nicotinic beta 1 subunit | 0.731 (104) | |
| LRRK2 | 0.0524 | | LRRK2 | leucine rich repeat kinase 2 | 0.998 (673) | |
| SLC29A1 | 0.0622 | | SLC29A1 | solute carrier family 29 member 1 (Augustine blood group) | 0.970 (104) | |
| PPP3CA | 0.0843 | | PPP3CA | protein phosphatase 3 catalytic subunit alpha | 0.994 (486) | |
| GRIN2D | 0.0921 | | GRIN2D | glutamate ionotropic receptor NMDA type subunit 2D | 0.846 (189) | |
| MPP2 | 0.0981 | | MPP2 | MAGUK p55 scaffold protein 2 | 0.331 (54) | |
| GRIN2B | 0.104 | | GRIN2B | glutamate ionotropic receptor NMDA type subunit 2B | 0.873 (252) | |
| MECP2 | 0.132 | | MECP2 | methyl-CpG binding protein 2 | 0.979 (398) | |
| ARRB2 | 0.136 | | ARRB2 | arrestin beta 2 | 0.980 (399) | |
| P2RX5 | 0.162 | | P2RX5 | purinergic receptor P2X 5 | 0.679 (127) | |
| MAPK8IP2 | 0.189 | | MAPK8IP2 | mitogen-activated protein kinase 8 interacting protein 2 | 0.723 (131) | |
| CHRNA7 | 0.205 | | CHRNA7 | cholinergic receptor nicotinic alpha 7 subunit | 0.920 (266) | |
| CHRNB2 | 0.206 | | CHRNB2 | cholinergic receptor nicotinic beta 2 subunit | 0.956 (244) | |
| RAB3GAP1 | 0.206 | | RAB3GAP1 | RAB3 GTPase activating protein catalytic subunit 1 | 0.949 (177) | |
| GRIN2A | 0.218 | | GRIN2A | glutamate ionotropic receptor NMDA type subunit 2A | 0.924 (331) | |
| TRPV1 | 0.235 | | TRPV1 | transient receptor potential cation channel subfamily V member 1 | 0.989 (340) | |
| CHRNA5 | 0.277 | | CHRNA5 | cholinergic receptor nicotinic alpha 5 subunit | 0.678 (102) | |
| OPRM1 | 0.299 | | OPRM1 | opioid receptor mu 1 | 0.978 (260) | |
| GSK3B | 0.302 | | GSK3B | glycogen synthase kinase 3 beta | 0.988 (448) | |
| CHRNE | 0.319 | | CHRNE | cholinergic receptor nicotinic epsilon subunit | 0.564 (83) | |
| SEZ6 | 0.321 | | SEZ6 | seizure related 6 homolog | 0.480 (58) | |
| GRIK5 | 0.363 | | GRIK5 | glutamate ionotropic receptor kainate type subunit 5 | 0.852 (180) | |
| GSK3A | 0.374 | | GSK3A | glycogen synthase kinase 3 alpha | 0.987 (453) | |
| SNCA | 0.376 | | SNCA | synuclein alpha | 0.998 (632) | |
| GRIN1 | 0.380 | | GRIN1 | glutamate ionotropic receptor NMDA type subunit 1 | 0.910 (304) | |
| ADORA1 | 0.488 | | ADORA1 | adenosine A1 receptor | 0.996 (394) | |
| S1PR2 | 0.505 | | S1PR2 | sphingosine-1-phosphate receptor 2 | 0.770 (126) | |
| GLRA3 | 0.507 | | GLRA3 | glycine receptor alpha 3 | 0.896 (116) | |
| CHRNA2 | 0.508 | | CHRNA2 | cholinergic receptor nicotinic alpha 2 subunit | 0.628 (90) | |
| NPFF | 0.539 | | NPFF | neuropeptide FF-amide peptide precursor | 0.327 (40) | |
| P2RX2 | 0.547 | | P2RX2 | purinergic receptor P2X 2 | 0.834 (176) | |
| DRD2 | 0.578 | | DRD2 | dopamine receptor D2 | 0.999 (619) | |
| GRID2 | 0.587 | | GRID2 | glutamate ionotropic receptor delta type subunit 2 | 0.830 (161) | |
| GHRL | 0.595 | | GHRL | ghrelin and obestatin prepropeptide | 0.999 (377) | |
| CDK5 | 0.645 | | CDK5 | cyclin dependent kinase 5 | 0.990 (580) | |
| AKT1 | 0.718 | | AKT1 | AKT serine/threonine kinase 1 | 0.997 (725) | |
| P2RX7 | 0.839 | | P2RX7 | purinergic receptor P2X 7 | 0.996 (571) | |
| GLRB | 0.895 | | GLRB | glycine receptor beta | 0.926 (141) | |
| CELF4 | 0.910 | | CELF4 | CUGBP Elav-like family member 4 | 0.743 (148) | |
| CHRNA4 | 0.930 | | CHRNA4 | cholinergic receptor nicotinic alpha 4 subunit | 0.764 (141) | |
| CHRNA3 | 0.960 | | CHRNA3 | cholinergic receptor nicotinic alpha 3 subunit | 0.890 (184) | |
| GRIK2 | 0.971 | | GRIK2 | glutamate ionotropic receptor kainate type subunit 2 | 0.928 (170) | |

Figure SF3: Details of gene ontology term “GO:0060079-Excitatory Postsynaptic Potential”

Table ST1A: Demographics of subjects.

| | Controls (CON) | Suicide Deaths (Sui) | p-value for statistical difference |
|---|---|---|---|
| N | 28 | 22 | N/A |
| Sex | Male= 13, Female=15 | Male= 12, Female=10 | p=0.57 |
| Age (Mean, Std. Dev) | 41.96 (15.41) | 39.73 (15.40) | p=0.61 |
| RIN (Mean, Std. Dev), CA1 Tissues | 5.41 (1.36) | 5.43 (1.65) | p=0.97 |
| PMI (Mean, Std. Dev) | 17.41 (6.99) | 21.60 (7.02) | *p=0.04 |
| RIN, iPSC-derived NPC | 9.5, 9.6 | 9.7, 9.8 | N/A |
| RIN, iPSC-derived Neurons | 10, 10 | 9.9, 10 | N/A |
| RIN=RNA Integrity Number, PMI=Postmortem Interval, N/A=Not Applicable. Unpaired t-test for age, RIN, and PMI. Chi-square test for sex differences. | | | |

Table ST1B: Detailed Demographics of Subjects.

| Sex | RIN | PMI (Hours) | Manner of Death | Comorbidity | Toxicology |
|---|---|---|---|---|---|
| CON Group | | | | | |
| M | 5.6 | 11 | Natural | None Reported | None |
| M | 3.1 | 19 | Natural | Head Injury | Ethanol |
| M | 3.8 | 17 | Accident | None Reported | None |
| M | 3.2 | 22 | Natural | None Reported | None |
| M | 6.2 | 28.35 | Natural | Tremors | None |
| M | 5.2 | 18 | Accident | None Reported | None |
| M | 5.9 | 15 | Accident | None Reported | None |
| M | 6.5 | 11.6 | Accident | None Reported | Ethanol |
| M | 3.1 | 17.36 | Natural | None Reported | None |
| F | 6.9 | 11.22 | Accident | Mild Stroke | Ethanol, Caffeine, Cotinine, Naloxone |
| F | 7.7 | 9.26 | Accident | None Reported | Caffeine, Cotinine, Benzoylecgonine, Codeine, Morphine, Diphenhydramine, 6-MAM |
| F | 4.8 | 8.2 | Accident | None Reported | Ethanol |
| F | 6.2 | 30.49 | Natural | None Reported | Midazolam |

| | | | | | |
|---|---|---|---|---|---|
| F | 6.4 | 27.16 | Undetermined | None Reported | Caffeine, Cotinine, Ethanol |
| F | 5.2 | 14.55 | Natural | Delusion | Cyclobenzaprine, 7-Aminoclonazepam |
| F | 5.5 | 17.3 | Homicide | None Reported | Caffeine |
| F | 3.2 | 24.1 | Natural | None Reported | None |
| F | 2.5 | 28.2 | Natural | None Reported | None |
| F | 5.5 | 22.59 | Accident | None Reported | None |
| F | 7.1 | 25.23 | Accident | None Reported | Caffeine, Methadone, Citalopram, EDDP, Amphetamine, Methamphetamine |
| F | 6.4 | 20 | Accident | None Reported | Lorazepam, 7-Amino Clonazepam, 11-hydroxy delta-9 THC, delta-9 carboxy THC, delta-9 THC, Amphetamine, Methamphetamine |
| M | 6.1 | 8 | Natural | None Reported | None |
| F | 5.6 | 11 | Accident | None Reported | Amphetamine, Methamphetamine |
| M | 6.3 | 18 | Accident | None Reported | THC |
| M | 5.9 | 6 | Accident | None Reported | Amphetamine, Methamphetamine, 6-Monoacetyl Morphine, Codeine, Morphine, Benzoylecgonine |
| F | 5.4 | 13 | Accident | None Reported | Amphetamine, Methamphetamine, Morphine, 6-Monoacetyl Morphine |
| M | 6.2 | 10 | Natural | Depression, Anxiety | Ethanol, Caffeine, Isopropanol, Cotinine, Naloxone |
| F | 6.1 | 24 | Natural | Lupus, Chronic Pain | Caffeine, Alprazolam, Delta-9 Carboxy THC, Trazodone, mCPP, Citalopram |
| Sui Group | | | | | |
| M | 3.9 | 16 | Suicide | Depression | Ethanol |
| M | 5.1 | 24 | Suicide | Suicide Ideation and Threats | Ethanol |
| M | 6.3 | 25 | Suicide | None Reported | None |
| M | 6.6 | 22.42 | Suicide | Depression, Anxiety | Trazodone, 7-Aminoclonazepam |
| M | 7.1 | 18 | Suicide | None Reported | None |
| M | 6.7 | 18.58 | Suicide | ADHD, Depression, Suicide Ideation | Amitryptyline |
| M | 4.2 | 15.17 | Suicide | None Reported | Ethanol |
| M | 6.9 | 16.51 | Suicide | None Reported | None |
| F | 6.3 | 24 | Suicide | None Reported | None |
| F | 6.8 | 43.28 | Suicide | None Reported | THC, 7-Amino Clonazepam |

| | | | | | |
|---|---|---|---|---|---|
| F | 5.5 | 18 | Suicide | None Reported | Caffeine, Cotinine, Diphenhydramine |
| F | 2.4 | 18.35 | Suicide | None Reported | Ethanol, Hydrocodone, Hydromorphine |
| F | 6.6 | 28 | Suicide | MDD | N/A |
| F | 2 | 27 | Suicide | MDD | N/A |
| F | 2.1 | 17 | Suicide | MDD | N/A |
| F | 4 | 22 | Suicide | MDD | N/A |
| M | 6.7 | 14 | Suicide | None Reported | Fluoxetine, Norfluoxetine |
| M | 5.9 | 9 | Suicide | None Reported | None |
| F | 6.9 | 27.41 | Suicide | OCD, Alcohol and Opioid Dependence | N/A |
| M | 6.8 | 24.47 | Suicide | MDD, ADHD | N/A |
| F | 4.8 | 28 | Suicide | MDD, OCD | N/A |
| M | 5.9 | 19 | Suicide | ASD, BD | N/A |

ADHD=Attention Deficit/Hyperactivity Disorder; MDD=Major Depressive Disorder; OCD=Obsessive Compulsive Disorder; ASD=Autism Spectrum Disorder; BD=Bipolar Disorder, N/A=Not Available. Green-highlighted Subjects= iPSCs were generated from the dermal fibroblasts of these subjects.

Table ST2: Significant gene ontology (GO) terms (CorrectedPvalue≤0.05; CorrectedMFPvalue≤0.05) obtained through gene score resampling (GSR) method of ErmineJ (version 3.2) using the p-value from DESeq2 results (factor:~age+sex+RIN+group). ID=numerical ID of the GO term; Genes# =number of the genes in the GO term; RawScore=ErmineJ-derived score for the GO term; Pval=uncorrected p-value; CorrPvalue=FDR-corrected p-value; MFPvalue=multifunctionality p-value, CorrMFPvalue=p-value after multifunctionality correction; Multifunc=multifunctionality; Green=NPAS4-associated GO terms; Orange=ALDH1A2-associated GO terms; Blue=ALDH1A2- and NAAA-associated GO terms.

| Name | ID | Genes # | RawScore | Pval | CorrPvalue | MFPvalue | CorrMFPvalue | Multifunc |
|---|---|---|---|---|---|---|---|---|
| Positive regulation of interferon-beta production | GO:0032728 | 38 | 1.21907837 | 1.00E-12 | 1.29E-09 | 3.89E-09 | 1.68E-06 | 0.802 |
| Regulation of interferon-beta production | GO:0032648 | 55 | 1.09966104 | 1.43E-12 | 1.54E-09 | 3.10E-07 | 9.11E-05 | 0.843 |
| Positive regulation of type I interferon production | GO:0032481 | 58 | 1.1062244 | 1.00E-12 | 2.15E-09 | 1.02E-06 | 2.44E-04 | 0.871 |
| Negative regulation of viral genome replication | GO:0045071 | 44 | 1.15847562 | 2.58E-12 | 2.38E-09 | 1.44E-10 | 7.74E-08 | 0.702 |
| Regulation of type I interferon production | GO:0032479 | 94 | 0.98294724 | 1.00E-12 | 3.23E-09 | 4.22E-06 | 9.10E-04 | 0.96 |
| Pattern recognition receptor signaling pathway | GO:0002221 | 84 | 1.00485783 | 1.00E-12 | 6.46E-09 | 1.05E-04 | 1.38E-02 | 0.973 |
| Toll-like receptor signaling pathway | GO:0002224 | 55 | 1.07259199 | 1.70E-11 | 1.37E-08 | 2.54E-04 | 2.78E-02 | 0.868 |
| Negative regulation of viral process | GO:0048525 | 73 | 0.98206521 | 1.09E-10 | 7.80E-08 | 3.35E-07 | 9.41E-05 | 0.814 |
| Regulation of viral genome replication | GO:0045069 | 69 | 0.91425383 | 5.69E-08 | 2.62E-05 | 4.27E-06 | 8.90E-04 | 0.836 |
| Cellular response to xenobiotic stimulus | GO:0071466 | 54 | 0.91800513 | 2.19E-06 | 7.09E-04 | 2.27E-06 | 5.06E-04 | 0.898 |
| Positive regulation of endothelial cell proliferation | GO:0001938 | 81 | 0.83095155 | 9.61E-06 | 2.59E-03 | 1.66E-05 | 2.91E-03 | 0.913 |
| Digestive tract development | GO:0048565 | 40 | 0.9073861 | 3.71E-05 | 6.15E-03 | 1.00E-12 | 1.08E-09 | 0.887 |
| Platelet activation | GO:0030168 | 48 | 0.87666977 | 4.79E-05 | 7.20E-03 | 1.78E-05 | 3.03E-03 | 0.839 |
| Excitatory postsynaptic potential | GO:0060079 | 49 | 0.87707 | 4.69E-05 | 7.22E-03 | 5.83E-10 | 2.69E-07 | 0.89 |
| Retinal metabolic process | GO:0042574 | 17 | 1.24282696 | 5.50E-05 | 8.08E-03 | 1.50E-05 | 2.69E-03 | 0.684 |
| Cellular response to retinoic acid | GO:0071300 | 51 | 0.86224105 | 5.72E-05 | 8.22E-03 | 1.85E-10 | 9.20E-08 | 0.87 |
| Midgut development | GO:0007494 | 6 | 1.87467553 | 6.00E-05 | 8.43E-03 | 7.50E-05 | 1.08E-02 | 0.744 |
| Embryonic camera-type eye development | GO:0031076 | 11 | 1.36407656 | 1.75E-04 | 1.66E-02 | 4.00E-05 | 6.01E-03 | 0.608 |
| Regulation of postsynaptic membrane potential | GO:0060078 | 76 | 0.7837523 | 2.31E-04 | 2.05E-02 | 1.54E-07 | 4.73E-05 | 0.906 |
| Embryonic digestive tract development | GO:0048566 | 14 | 1.23906795 | 2.30E-04 | 2.06E-02 | 1.90E-04 | 2.19E-02 | 0.731 |
| Response to retinoic acid | GO:0032526 | 88 | 0.76959343 | 2.60E-04 | 2.16E-02 | 1.61E-06 | 3.70E-04 | 0.969 |
| Long-term memory | GO:0007616 | 35 | 0.89977609 | 2.69E-04 | 2.20E-02 | 1.16E-05 | 2.21E-03 | 0.838 |
| Primary alcohol metabolic process | GO:0034308 | 62 | 0.80199472 | 3.66E-04 | 2.57E-02 | 1.00E-12 | 1.62E-09 | 0.9 |
| Proximal/distal pattern formation | GO:0009954 | 17 | 1.12669608 | 3.90E-04 | 2.60E-02 | 1.25E-04 | 1.55E-02 | 0.723 |
| Embryonic forelimb morphogenesis | GO:0035115 | 21 | 1.05611122 | 4.70E-04 | 2.87E-02 | 2.20E-04 | 2.49E-02 | 0.784 |
| Retinoic acid biosynthetic process | GO:0002138 | 7 | 1.50579959 | 5.00E-04 | 2.99E-02 | 1.00E-12 | 3.23E-09 | 0.52 |
| Forelimb morphogenesis | GO:0035136 | 25 | 1.00171502 | 7.00E-04 | 3.74E-02 | 4.60E-04 | 4.87E-02 | 0.792 |
| Fc receptor signaling pathway | GO:0038093 | 44 | 0.84070362 | 7.31E-04 | 3.84E-02 | 9.06E-05 | 1.25E-02 | 0.883 |
| Vitamin A metabolic process | GO:0006776 | 7 | 1.4515446 | 7.60E-04 | 3.93E-02 | 1.00E-12 | 2.15E-09 | 0.471 |

Table ST3: DESeq2 results for Differentially Expressed Genes (DEGs) in iPSC-derived hippocampal-like neurons (NPC vs Neurons). n=4 for both the NPCs and Neurons.

| Gene symbol | P-value | FDR step up | Ratio | Fold change | LSMean (NPC) | LSMean (Neurons) | Change in Neurons |
|---|---|---|---|---|---|---|---|
| Neuron Markers | | | | | | | |
| MAP2 | 1.22E-10 | 7.27E-10 | 0.50 | -2.00 | 20216.47 | 40372.30 | Increased |
| DLG4 | 2.08E-13 | 1.73E-12 | 0.33 | -3.07 | 1092.41 | 3356.51 | Increased |
| TUBB3 | 7.76E-05 | 2.09E-04 | 0.46 | -2.18 | 10699.69 | 23301.64 | Increased |
| Hippocampal Markers | | | | | | | |
| ZBTB20 | 4.64E-06 | 1.50E-05 | 0.55 | -1.82 | 2258.07 | 4102.88 | Increased |
| LHX9 | 2.65E-02 | 4.42E-02 | 0.11 | -8.70 | 99.49 | 865.57 | Increased |
| ELAVL2 | 4.86E-05 | 1.35E-04 | 0.37 | -2.72 | 1555.09 | 4223.59 | Increased |
| TSPAN7 | 2.23E-31 | 1.02E-29 | 0.09 | -10.67 | 341.84 | 3647.89 | Increased |
| CA1 Markers | | | | | | | |
| POU3F1 | 2.45E-11 | 1.58E-10 | 3.72 | 3.72 | 671.81 | 180.51 | Decreased |
| BCL11B | 1.51E-04 | 3.88E-04 | 0.24 | -4.17 | 121.61 | 506.50 | Increased |
| WFS1 | 3.13E-01 | 3.88E-01 | 1.16 | 1.16 | 823.45 | 710.76 | Unchanged |
| CA2 Markers | | | | | | | |
| PCP4 | 1.94E-26 | 5.78E-25 | 0.06 | -17.20 | 21.92 | 376.90 | Increased |
| RGS14 | 4.78E-05 | 1.33E-04 | 2.71 | 2.71 | 310.55 | 114.57 | Decreased |
| CA3 Markers | | | | | | | |
| ELAVL2 | 4.86E-05 | 1.35E-04 | 0.37 | -2.72 | 1555.09 | 4223.59 | Increased |
| GRIK4 | 7.24E-24 | 1.71E-22 | 0.19 | -5.15 | 97.73 | 503.64 | Increased |
| SNAP91 | 1.25E-14 | 1.19E-13 | 0.08 | -12.22 | 182.29 | 2228.27 | Increased |
| SH3GL2 | 6.26E-26 | 1.77E-24 | 0.05 | -20.24 | 73.48 | 1487.06 | Increased |
| CADPS2 | 2.38E-10 | 1.37E-09 | 0.40 | -2.50 | 403.03 | 1007.88 | Increased |
| CHGB | 1.18E-15 | 1.28E-14 | 0.20 | -5.01 | 314.53 | 1575.08 | Increased |
| CACNA1A | 3.44E-07 | 1.32E-06 | 0.32 | -3.16 | 102.71 | 324.72 | Increased |
| LPL | 1.30E-19 | 2.06E-18 | 16.51 | 16.51 | 23864.67 | 1445.89 | Decreased |
| TGFB2 | 2.82E-04 | 6.87E-04 | 0.15 | -6.55 | 470.78 | 3082.20 | Increased |
| MYO5B | 1.31E-11 | 8.74E-11 | 0.35 | -2.88 | 363.62 | 1046.40 | Increased |
| PKIA | 5.83E-12 | 4.03E-11 | 0.29 | -3.40 | 2909.73 | 9897.58 | Increased |
| SNCA | 1.29E-06 | 4.52E-06 | 0.38 | -2.62 | 749.64 | 1963.01 | Increased |
| SPOCK1 | 6.84E-12 | 4.69E-11 | 2.94 | 2.94 | 5093.39 | 1732.34 | Decreased |
| HSPA2 | 6.78E-04 | 1.54E-03 | 0.36 | -2.75 | 301.57 | 829.65 | Increased |
| NEFL | 2.39E-03 | 4.91E-03 | 0.60 | -1.66 | 7239.41 | 12038.06 | Increased |
| DKK3 | 3.68E-30 | 1.58E-28 | 0.23 | -4.26 | 1705.69 | 7268.11 | Increased |
| DG Markers | | | | | | | |
| PROX1 | 8.00E-02 | 1.18E-01 | 1.63 | 1.63 | 743.05 | 454.89 | Unchanged |

| | | | | | | | |
|---|---|---|---|---|---|---|---|
| CALB1 | 5.27E-08 | 2.25E-07 | 5.55 | 5.55 | 6930.12 | 1248.18 | Decreased |
| DOCK10 | 9.88E-12 | 6.66E-11 | 0.21 | -4.81 | 475.82 | 2286.60 | Increased |

Table ST4: Differentially expressed genes (DEGs), determined by RNA-Sequencing (RNA-Seq), in iPSC-derived hippocampal-like NPCs of suicide deaths (CON vs Sui). n=2/group. Highlighted genes are common in both NPCs and neurons.

| DESeq2 results for hippocampal-like NPCs (CON vs Sui), p≤0.05, FDR≤0.05 | | | | | | |
|---|---|---|---|---|---|---|
| Gene Symbol | P-value | FDR step up | Ratio | Fold change | LSMean(CON) | LSMean(Sui) |
| *SNORD3B-2* | 5.29E-88 | 3.22E-84 | 0.02 | -57.59 | 68.08 | 3920.73 |
| *SNORD3B-1* | 5.29E-88 | 3.22E-84 | 0.02 | -57.59 | 68.08 | 3920.73 |
| *RELN* | 1.04E-12 | 4.23E-09 | 5.55 | 5.55 | 3215.45 | 579.00 |
| *CRH* | 8.39E-10 | 2.56E-06 | 5.00 | 5.00 | 866.32 | 173.37 |
| *CNTN2* | 3.78E-09 | 9.22E-06 | 4.23 | 4.23 | 3894.81 | 920.86 |
| *H4C11* | 2.68E-08 | 5.44E-05 | 0.29 | -3.43 | 900.78 | 3092.58 |
| *SCUBE1* | 8.21E-08 | 1.43E-04 | 3.31 | 3.31 | 3370.64 | 1019.14 |
| *EMX2OS* | 3.19E-07 | 4.86E-04 | 4.28 | 4.28 | 1074.58 | 251.21 |
| *PARM1* | 6.48E-07 | 8.77E-04 | 2.91 | 2.91 | 2925.97 | 1004.88 |
| *TMTC1* | 2.13E-06 | 2.60E-03 | 2.51 | 2.51 | 3019.33 | 1201.43 |
| *COL3A1* | 3.03E-06 | 3.36E-03 | 0.50 | -2.00 | 8849.73 | 17657.70 |
| *HMCN1* | 5.56E-06 | 5.52E-03 | 0.36 | -2.76 | 597.42 | 1649.41 |
| *CNTNAP2* | 5.89E-06 | 5.52E-03 | 2.14 | 2.14 | 5905.46 | 2764.22 |
| *EMX2* | 7.54E-06 | 6.57E-03 | 4.24 | 4.24 | 587.40 | 138.49 |
| *SCG2* | 1.17E-05 | 9.50E-03 | 2.74 | 2.74 | 9006.52 | 3285.05 |
| *OXTR* | 2.49E-05 | 1.90E-02 | 3.41 | 3.41 | 515.49 | 151.01 |
| *PDPR* | 3.30E-05 | 2.20E-02 | 0.46 | -2.16 | 1664.37 | 3588.05 |
| *H2AC12* | 3.65E-05 | 2.20E-02 | 0.53 | -1.87 | 4910.13 | 9206.07 |
| *COL8A1* | 3.66E-05 | 2.20E-02 | 2.70 | 2.70 | 3610.26 | 1339.34 |
| *BGN* | 3.77E-05 | 2.20E-02 | 0.34 | -2.93 | 255.28 | 748.74 |
| *CCZ1* | 3.79E-05 | 2.20E-02 | 2.23 | 2.23 | 2006.89 | 901.72 |
| *BLACAT1* | 4.06E-05 | 2.25E-02 | 3.49 | 3.49 | 688.16 | 197.13 |
| *PRSS23* | 5.16E-05 | 2.65E-02 | 1.85 | 1.85 | 13209.09 | 7147.09 |
| *FBLN2* | 5.40E-05 | 2.65E-02 | 0.45 | -2.24 | 1125.37 | 2523.00 |
| *ABCA1* | 5.43E-05 | 2.65E-02 | 2.23 | 2.23 | 1888.39 | 847.99 |
| *FZD5* | 5.82E-05 | 2.73E-02 | 2.58 | 2.58 | 809.60 | 313.30 |
| *IFITM2* | 6.19E-05 | 2.78E-02 | 0.38 | -2.62 | 793.15 | 2077.36 |
| *UNC5D* | 6.39E-05 | 2.78E-02 | 2.24 | 2.24 | 1654.83 | 738.10 |
| *CCZ1B* | 7.79E-05 | 3.28E-02 | 0.49 | -2.06 | 1171.02 | 2407.12 |
| *COL12A1* | 9.57E-05 | 3.89E-02 | 0.39 | -2.57 | 1172.34 | 3017.78 |
| *VAV3* | 1.10E-04 | 4.32E-02 | 2.39 | 2.39 | 1332.99 | 557.65 |

Table ST5: Differentially expressed genes (DEGs), determined by RNA-Sequencing (RNA-Seq), in iPSC-derived hippocampal-like Neurons of suicide deaths (Con vs Sui). n=2/group. Highlighted genes were common in both NPCs and neurons.

| DESEq2 results for hippocampal-like neurons (CON vs Sui), p≤0.05, FDR≤0.05 | | | | | | |
|---|---|---|---|---|---|---|
| Gene symbol | P-value | FDR step up | Ratio | Fold change | LSMean(CON) | LSMean(Sui) |
| *SNORD3B-2* | 8.32E-31 | 5.89E-27 | 0.02 | -40.71 | 35.19 | 1432.48 |
| *SNORD3B-1* | 8.32E-31 | 5.89E-27 | 0.02 | -40.71 | 35.19 | 1432.48 |
| *COL3A1* | 1.11E-15 | 5.23E-12 | 0.22 | -4.50 | 8656.00 | 38989.35 |
| *COL1A1* | 1.69E-15 | 5.97E-12 | 0.13 | -7.51 | 2230.48 | 16757.22 |
| *DCN* | 4.03E-13 | 1.14E-09 | 0.21 | -4.86 | 411.79 | 2003.27 |
| *IGFBP3* | 1.57E-11 | 3.70E-08 | 0.20 | -5.12 | 229.47 | 1175.62 |
| *COL5A1* | 2.42E-11 | 4.89E-08 | 0.24 | -4.22 | 944.15 | 3982.05 |
| *HMCN1* | 8.95E-11 | 1.54E-07 | 0.16 | -6.15 | 113.60 | 698.99 |
| *THBS1* | 9.78E-11 | 1.54E-07 | 0.23 | -4.28 | 525.45 | 2251.32 |
| *LUM* | 1.65E-10 | 2.33E-07 | 0.28 | -3.57 | 1128.19 | 4029.03 |
| *COL6A3* | 1.96E-10 | 2.52E-07 | 0.18 | -5.60 | 942.49 | 5279.06 |
| *H4C11* | 4.95E-08 | 5.83E-05 | 0.18 | -5.57 | 197.46 | 1099.07 |
| *PRRX1* | 1.14E-07 | 1.18E-04 | 0.25 | -4.02 | 168.74 | 678.51 |
| *SNORD97* | 1.23E-07 | 1.18E-04 | 0.27 | -3.67 | 315.25 | 1156.59 |
| *IFITM2* | 1.25E-07 | 1.18E-04 | 0.23 | -4.27 | 267.81 | 1144.73 |
| *FBN2* | 2.65E-07 | 2.35E-04 | 0.26 | -3.83 | 499.27 | 1914.45 |
| *ID1* | 3.70E-07 | 3.08E-04 | 0.27 | -3.75 | 181.85 | 682.55 |
| *POSTN* | 5.68E-07 | 4.46E-04 | 0.21 | -4.81 | 697.52 | 3351.59 |
| *CYP26A1* | 6.00E-07 | 4.46E-04 | 0.21 | -4.72 | 100.56 | 474.93 |
| *NOMO3* | 7.90E-07 | 5.59E-04 | 0.20 | -5.09 | 481.99 | 2455.69 |
| *FBN1* | 1.99E-06 | 1.34E-03 | 0.35 | -2.86 | 745.52 | 2134.39 |
| *MME* | 4.39E-06 | 2.83E-03 | 0.17 | -5.99 | 48.15 | 288.61 |
| *MMP2* | 5.46E-06 | 3.36E-03 | 0.25 | -4.01 | 184.15 | 738.88 |
| *PDGFRA* | 6.62E-06 | 3.90E-03 | 0.33 | -3.07 | 283.64 | 870.61 |
| *MGP* | 9.81E-06 | 5.55E-03 | 0.21 | -4.86 | 69.97 | 339.72 |
| *ALDH1A3* | 2.90E-05 | 1.58E-02 | 0.15 | -6.64 | 59.00 | 391.60 |
| *COL5A2* | 3.90E-05 | 2.04E-02 | 0.44 | -2.28 | 3432.70 | 7821.19 |
| *AQP1* | 4.55E-05 | 2.30E-02 | 0.33 | -2.99 | 396.99 | 1186.18 |
| *CCZ1B* | 5.12E-05 | 2.50E-02 | 0.42 | -2.41 | 720.36 | 1734.82 |
| *CXCL14* | 9.87E-05 | 4.65E-02 | 0.14 | -7.23 | 19.88 | 143.68 |
| *NLRP2* | 1.02E-04 | 4.65E-02 | 0.28 | -3.58 | 117.21 | 419.51 |
| *MXRA5* | 1.05E-04 | 4.66E-02 | 0.23 | -4.31 | 73.19 | 315.56 |
| *TBR1* | 1.11E-04 | 4.78E-02 | 6.39 | 6.39 | 165.84 | 25.95 |

Materials and Methods

RNA Extraction and Sequencing

The postmortem CA1 tissues were dissected out from the hippocampal formation and stored in RNAlater solution (Thermofisher # AM7020) at -80°C until RNA extraction. The total RNA was extracted from CA1 tissues using RNeasy Lipid Tissue Mini Kit (Qiagen # 74804) as per manufacturer's instructions. The total RNA from iPSC-derived NPCs and neurons were extracted using RNeasy Mini Kit (Qiagen # 74104). The RNA library preparation was carried out using Ribo-Zero Gold rRNA depletion and TrueSeq Stranded Total RNA Kit (Illumina, San Diego, USA). For 36 samples, 100 bp paired-end sequencing was performed on NovaSeq 6000 platform at the UCI Genomics High Throughput Facility (GHTF). For 14 samples, 125 bp paired-end sequencing was performed on HiSeq 2500 platform at the University of Utah High-Throughput Genomics. The number of read for each sample ranged from 27,277,523 to 65,709,547.

Gene-level Differential Expression (DE) and Gene Ontology (GO) Analyses

Differentially expressed genes (DEGs) analysis was performed in Partek Flow, following five (5) steps: (1) rDNA and mtrDNA contaminants were filtered out from raw fastq files through Bowtie 2 (version 2.2.5). (2) Adaptor sequences were trimmed through Cutadapt-1.12. (3) Alignment of the fastq files was performed through TopHat 2 (version 2.1.0) with default settings [assembly: human-hg19, index: whole genome (administrator)]. (4) Annotation was performed with task "Quantify to annotation model (Partek E/M)" with assembly of human-hg19 and annotation model of RefSeq Transcript 93-2020-02-03. (5) Finally, differentially expressed genes (DEGs) were determined through DESeq2 with default settings. For CA1 tissue DEG analysis, features with minimum value of zero (0) was excluded before DESeq2 analysis. DESeq2 was run with factor of ~Age+Sex+RIN+Group with default parameters. For DEG analysis in iPSC-derived hippocampal-like NPCs and neurons, features with minimum value of ≤10 were excluded from analysis before DESeq2. DESeq2 was run with default parameters with factors of ~Age+Group (RIN was excluded because RIN was similar in all samples) in CON vs Sui comparison while factor of ~Age+Sex+Type in NPC vs Neurons comparison. The gene ontology (GO) analysis was performed using the gene score resampling (GSR) method as implemented in the ErmineJ (Gillis, Mistry, & Pavlidis, 2010). GSR does not require a threshold for gene selection, but rather uses all the genes with their p-values. The full list of genes with their p-value from the DESeq2 (factor: ~age+sex+RIN+group) was used in the analysis. The following settings were used in the GSR analysis: aspect=biological process, maximum gene set size=100, minimum gene set size=5, maximum iterations=200,000.

Dendritic Spine Morphology Analyses

Briefly, freshly dissected out hippocampal blocks (~1$cm^3$) were immersion fixed in freshly prepared cold 4% paraformaldehyde (in 0.1M Phosphate buffer (PB)) for 1 hour. After 1 hour of fixation, hippocampal blocks were post-fixed in 4% paraformaldehyde + 0.125% glutaraldehyde for 24 hours at 4°C. Later, 350 µm coronal slices were obtained through vibratome (Leica VT1200). Microscopic 1,1'-dioctadecyl-3,3,3',3'-tetramethylindocarbocyanine perchlorate (DiI) crystals (Thermofisher # D282) were placed in CA1 regions and incubated in 0.1M PB at 37°C for 72 hours. Dendritic spine images were obtained through a Zeiss LSM 710 confocal microscope using a 63 x oil immersion objective (NA=1.4). The acquired images of dendritic spines were deconvolved using AutoQuant X3 (Bitplane, RRID:SCR_002465) and three-dimensional dendritic spine modeling was performed using Neurolucida 360 (MBF Bioscience, RRID: SCR_016788). 2-3 dendritic segments were analyzed per neuron for both apical and basal dendrites.

Conversion of Skin Fibroblasts into iPSC:

Briefly, dermal fibroblasts were grown from skin biopsies and maintained in fibroblasts media [ DMEM, FBS (10%), MEM NEAA, Na-Pyruvate, GlutaMAX, Penicillin-Streptomycin and beta-mercaptoethanol]. Fibroblasts (in fibroblasts media) were transfected with three episomal plasmids expressing human OCT3/4 and shRNA against p53 (Addgene# 27077: pCXLE-hOCT3/4-shp53-F), L—MYC and LIN28 (Addgene# 27080: pCXLE-hUL), SOX2 and KLF4 (Addgene# 27078: pCXLE-hSK). Fibroblasts ($1x10^6$ cells, in fibroblasts media), less than 5 passages, were electroporated with 3 µg of each episomal plasmid through Neon™ Transfection System 100 µL kit (MPK10096, Thermo Fisher Scientific, MA, USA) according to manufacturer protocol. Neon™ transfection conditions were 1650 V, 10 ms and 3 pulses. Seven-days post-transfection, fibroblasts were replated onto 6-well plates containing mitomycin C-inactivated mouse embryonic fibroblasts (MEF) feeder cells. Next day, fibroblasts media was switched to iPSC media [ DMEM/F12 with 20% KOSR, 3% ESC-quality FBS, 20 ng/ml bFGF, 1mM GlutaMAX, 100 µM MEM NEAA, 100 µM 2-mercaptoethanol, and penicillin-streptomycin] which was changed every day until iPSC colonies were ready to be picked. Colonies were manually picked 21 days post-transfection and maintained on Vitronectin XF (Stemcell Technologies Inc, #07180) coated wells with mTeSR Plus feeder-free media (Stemcell Technologies Inc, #100-0276). Pluripotency of iPSCs were characterized through immunostaining of pluripotency markers OCT4, SOX2, SSEA4 and TRA-1-60 (A24881, Thermo Fisher Scientific, MA, USA).

Generation of Hippocampal-like Neural Precursor Cell (NPC) and Neurons:

Briefly, iPSCs were mechanically dissociated and transferred to low-attachment cell culture plates with mTeSR Plus media (with the addition of ROCK inhibitor, 10 µM) to form embryoid body (EB). After two days, mTeSR Plus media was replaced with N2/B27 media [DMEM/F12/GlutaMax/N2/B27(w/o vitamin A) /Penstrep] supplemented with DKK1 (0.5 µg/mL), SB431542 (10 µM), Noggin (0.5 µg/mL) and cyclopamine (1 µM) (maintained for 10 days). After 10 days, EBs were transferred onto poly-ornithine/laminin coated plates with N2/B27 media plus laminin (1 µg/mL). Once rosettes formation was visible, EBs were mechanically dispersed and maintained in N2/B27 media supplemented with 20 ng/mL human basic fibroblasts growth factor (b-FGF) to obtain hippocampal-like NPC. To obtain hippocampal-like neurons, NPCs were plated onto poly-ornithine/laminin coated plates and maintained in neuronal differentiation media [N2B27 media + ascorbic acid (200 µM) + dibutyryl cyclic-AMP (400 µM) + BDNF (20 ng/ML) and Wnt3A (5 ng/mL). Wnt3A was discontinued after three weeks and the differentiation was maintained for two more weeks. The iPSC-derived neurons were characterized through RNA-Seq and immunocytochemistry (ICC). The following primary antibodies were used:

MAP2 (ZRB2290, Sigma-Aldrich), beta-Tubulin-III (Tuj1) (801202, BioLegend), vGluT1 (821302, BioLegend), PSD95 (MA1-045, Thermofisher), GABA (A2052, Sigma-Aldrich), GRIK4 (NBP1-76852, Novus Biologicals), PROX1 (925202, BioLegend), and CTIP2 (650601, BioLegend).

References:

Gillis, J., Mistry, M., & Pavlidis, P. (2010). Gene function analysis in complex data sets using ErmineJ. *Nat Protoc, 5*(6), 1148-1159. doi:10.1038/nprot.2010.78
